# Client preferences for service delivery during the early treatment period in South Africa and Zambia: Mixed-method findings from a discrete choice experiment and concurrent focus group discussions

**DOI:** 10.1101/2025.07.28.25332159

**Authors:** Allison Morgan, Linda Sande, Mhairi Maskew, Sydney Rosen, Lawrence Long, Nyasha Mutanda, Sophie Pascoe, Caroline Govathson, Aniset Kamanga, Taurai Makwalu, Priscilla Lumano-Mulenga, Lufuno Malala, Musa Manganye, Prudence Haimbe, Hilda Shakwelele, Elizabeth Kachingwe, Thandiwe Ngoma, Nancy Scott

## Abstract

**Background:** Disengagement from antiretroviral therapy (ART) is common in the first 6 months of HIV treatment in sub-Saharan Africa. Using mixed-methods we aimed to understand preferences during this early treatment period.

**Methods:** Between 8/2023-11/2023, adults who had initiated/re-initiated ART a median of 8 months prior were enrolled at 18 healthcare facilities across South Africa (SA) and Zambia to participate in a discrete choice experiment (DCE) and focus group discussion (FGD). In the DCE, participants made 9 choices between unique service delivery scenarios (each comprised of 8 attributes). Analyzed using a conditional logit model, we report findings using odds ratios (95% confidence intervals). Following the DCE, FGDs explored barriers to care seeking and care preferences. Thematic analysis was used to interpret FGDs. DCE and FGD findings were triangulated to understand preferences.

**Results:** We enrolled 250 respondents: 128 in Zambia (55% female, median age 35); 122 in SA (83% female, median age 33). Community-based services were less favorable to respondents than clinic-based care (SA: 0.62 (95% CI 0.52, 0.75); Zambia: 0.44 (0.36, 0.53)). Respondents preferred 6-month dispensing (SA: 1.3 (1.1, 1.6); Zambia: 2.1 (1.8, 2.6)) to 1-month intervals. Respondents also preferred accessing services from friendly providers. Qualitative insights corroborated DCE findings. They also revealed frustrations with long wait times at clinics.

**Conclusion:** Utilizing a decision experiment with qualitative methods allowed us to uniquely capture drivers of client decision-making and the nuanced factors that shape experiences. Results suggest that enrollment in lower-intensity models may improve client experiences during the early treatment period.

## Introduction

Disengagement from antiretroviral therapy (ART) remains high among people living with HIV (PLHIV) in sub-Saharan Africa, slowing countries’ progress toward the global 95-95-95 targets [1]. Recent literature suggests attrition is especially high during the first 6 months on treatment, known as the early treatment period [2]. A large body of published literature has explored reasons for disengagement from care and client preferences [3], but very little of it examines as a distinct experience the early treatment period, when clients are still adjusting to their HIV diagnosis, becoming familiar with clinic environments and schedules, and coping with the logistical, social, and personal burdens of lifelong treatment. The preferences of ART clients during the early treatment period are poorly documented, hampering the development of models of service delivery aimed explicitly at the unique needs of newly initiating/re-initiating clients. While the service delivery landscape has changed in recent years with the introduction of patient-centered, low-intensity, differentiated service delivery (DSD) models [4], eligibility for these models has traditionally been limited to virally suppressed patients with at least 6 months’ experience on ART. Newly initiating or re-initiating clients continue to be offered a relatively intensive, “one size fits all” model of service delivery which does not necessarily reflect clients’ needs or preferences as they begin or return to treatment [2].

Originating in economics, discrete choice experiments (DCEs) are a quantitative stated-preference research method used to estimate the relative importance of specific attributes. This is done by asking respondents to choose their preferred option between various hypothetical scenarios, essentially allowing researchers to understand what is driving behavior and what tradeoffs people are willing to make, without needing to observe it [5,6]. Originating in sociology, focus group discussions (FGDs) are a qualitative research method used to explore group norms and collective meaning [7]. In global health, FGDs are frequently used to illuminate community perspectives, health behaviors, lived experience, and the social context [8]. Qualitative methods such as FGDs and DCEs are often used sequentially in mixed-methods research, with the qualitative data informing the design of a DCE, including attribute selection[9–13]. A review of the concurrent use of the two approaches, however, found that qualitative methods have been underutilized, or at least underreported, in the interpretation of healthcare-related DCE preference results [9]. That review argued for the value of using qualitative methods not just to design DCEs but to enrich the analysis and interpretation of preference results.

Within HIV cascade of care research, a growing number of published studies have used DCEs to understand HIV clients’ preferences for HIV prevention, testing, and service delivery [14–21], and many of these have used qualitative data methods to inform the design of the DCE [22]. None, however, has conducted the two types of data collection simultaneously and with the same participants [15]. Using these complementary data collection methods simultaneously offers the opportunity to explore patient preferences from different perspectives and to better understand complexities that shape decisions.

As part of a project that aims to identify clients’ preferences and experiences in the first 6 months on ART, we conducted a concurrent mixed methods study that used findings from a quantitative client survey to inform the design of a subsequent, joint DCE and FGDs [23]. This approach allowed us to understand the trade-offs that clients are willing to make in the early treatment period and the qualitative context and lived experience behind their decisions, strengthening our interpretations of client preferences. Together, these methods provided a more comprehensive picture of client priorities in the early treatment period and could inform the design of new service delivery models that are both evidence-based and client-centered.

## Methods

### Study overview and setting

The PREFER study [23] was a mixed-methods observational cohort that followed adults who had been on ART for six months or less at 18 facilities in South Africa’s Gauteng, KwaZulu Natal, and Mpumalanga provinces and 12 facilities in Zambia’s Central and Lusaka provinces between 7 September 2022 and 29 November 2023. It aimed to understand the needs and preferences of people starting or restarting HIV treatment to inform the design of differentiated service delivery (DSD) models, improve early treatment outcomes, and identify distinct challenges faced by naive clients compared to those returning to care [23,24]. PREFER started with a baseline, quantitative survey [24], which was followed by the DCE and FGDs reported here. The DCE and FGDs were conducted at a subset of 11 PREFER sites in South Africa between 21 August and 8 October 2023 and 7 sites in Zambia between 18 Aug and 13 September 2023. This subset of sites was purposively selected to balance urban and rural locations.

### Study population, recruitment, procedures, and sample size

DCE and FGD participants were selected from the pool of PREFER baseline survey participants. Participants were eligible for the DCE and FGD if, in the PREFER baseline survey, they reported at least one challenge with their care (missing a scheduled clinic visits by 2 days or more, anticipating difficulty adhering to or collecting medications, or feeling their clinical care was worse than expected). A systematic random selection of eligible participants was contacted approximately 6 months after PREFER survey enrollment and invited to participate in the DCE and FGD. Due to the design of the PREFER baseline quantitative survey, which enrolled clients with 0-6 months experience on ART, participants had spent varying amounts of time on treatment since initiation (6-11 months) when FGD and DCE data were collected.

We aimed to include 150 of the baseline survey respondents per country (300 in total) to participate in the DCE, of whom a maximum of 75 per country (150 in total) would also participate in a FGD (8-10 participants per group; maximum of 15 groups). This sample size was sufficient to reach saturation for the qualitative FGDs [25] and met the minimum sample necessary per stratum for DCE validity (per our study design, the minimum acceptable sample size was 94 participants) [26].

### Data collection and management

Eligible participants were asked to make a separate visit to the study site at a scheduled time to participate in the study activity. Upon arrival, eligibility was confirmed and informed consent administered. In Zambia, participants were separated by sex for study procedures, due to local preferences related to the sensitive nature of the topic; men and women participated jointly in South Africa as fewer men were enrolled in the baseline survey there. Once consented, participants individually completed the DCE survey on paper. FGDs were conducted immediately after completion of the DCE. Study team members were trained in qualitative research methods, the interview guide, administration of the DCE, and in human subject research ethics. Participants were given a transport reimbursement and lunch allowance equivalent to 170 Zambian Kwacha (USD $6.00) or 100 South African Rand (USD $5.40). Participant IDs were used to link responses with demographic and ART history data from the PREFER baseline survey. Study personnel later entered DCE data from paper forms into SurveyCTO (Dobility, Inc.) for storage and analysis.

### Discrete choice experiment design and analysis

This DCE was designed to identify which characteristics of ART service delivery were most important to a client’s care experience in the first six months and might influence their decision to remain in care during this period. A DCE does this by asking participants to make repeated choices between scenarios each described by various characteristics (attributes) set at different values (levels) [9,27]. We reported against the DIRECT Checklist for Discrete Choice Experiments in Health [28].

The attributes and levels were adapted from a DCE conducted by colleagues in a similar context [13]; contextual refinements were informed by the published literature [2] and the PREFER quantitative survey results [24]. Each attribute was selected because it was expected to affect the care-seeking experience and because it was potentially modifiable at clinics in South Africa and Zambia. Other variables often used in this context include additional healthcare provider characteristics, confidentiality of services, incentives and types of services offered. Due to the nature of our study population and the burden that the early treatment period poses, however, we decided to forego some of these additional variables to focus specifically on appointment scheduling and dispensing.

To ensure that participants can cognitively discern between scenarios, a maximum of 10 attributes and 2-3 levels per attribute is generally recommended for DCE studies [11,27]. We selected eight attributes of care service delivery, seven with three levels and one with two levels (Panel 1). This number of attributes and levels yields over 4,000 possible alternative scenarios, which is impossible to test using a full factorial design. We therefore used Stata’s Federov algorithm (StataCorp 2023) to generate a maximally efficient fractional factorial design consisting of 18 choice sets. We then assessed the efficient design to ensure that it was orthogonal and balanced [29] (See Supplementary File 1). Each choice set contained two alternative scenarios offering different aspects of care with contextually-relevant pictures depicting each attribute level (Figure 1).

**Figure 1.**
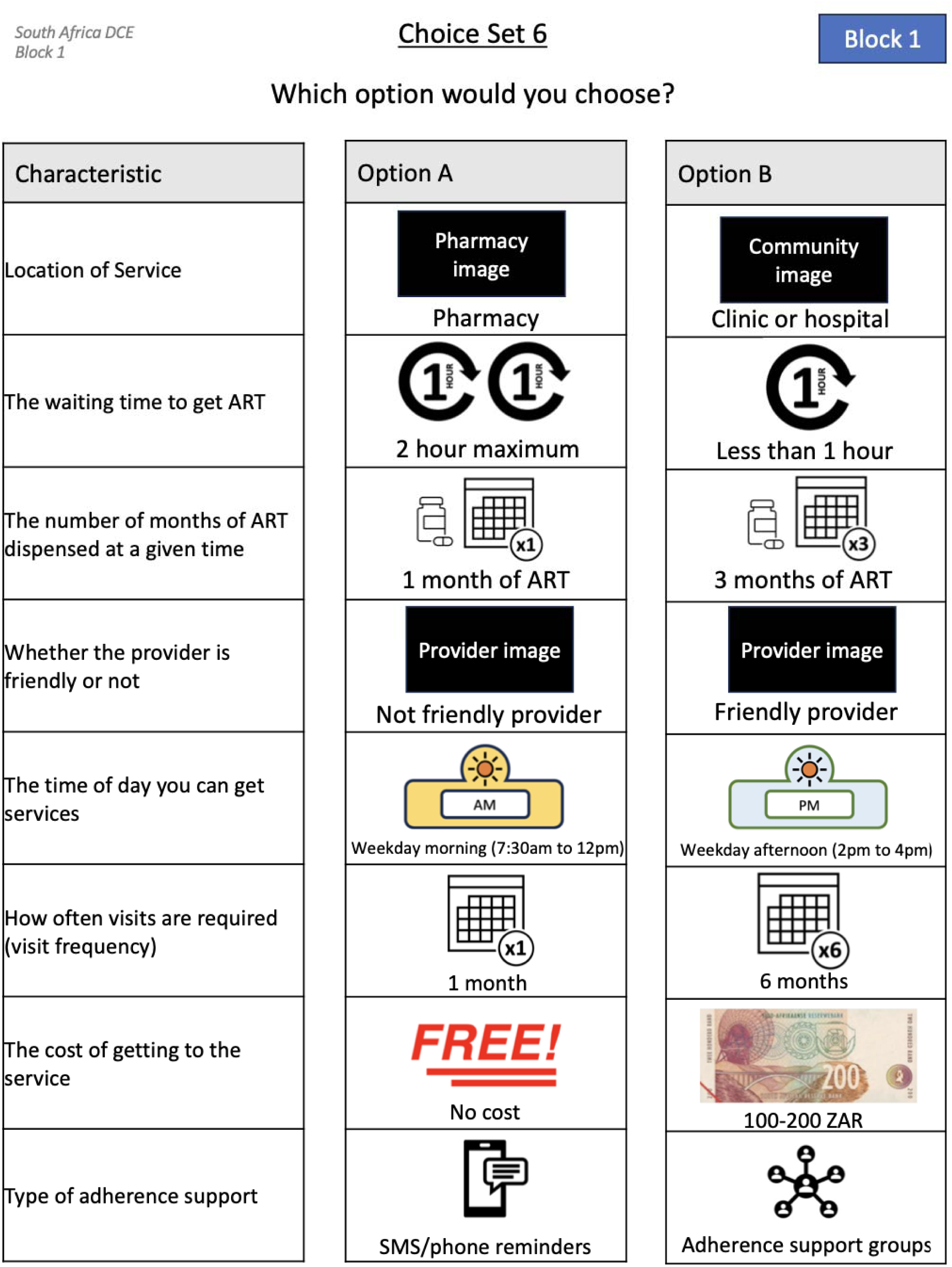
Example of PREFER DCE choice task.

The 18 choice sets were then split into two blocks of 9 choice sets each (Block 1 and Block 2). Each participant completed only one block of 9 choice sets to minimize cognitive fatigue [29]. All participants at each site completed the same block, with blocks alternating by site [29]. We used a forced choice approach, meaning that participants were required to select a scenario from each choice set; that is, they could not opt out or choose “none of the above”. This approach increases statistical efficiency and maximizes the information that can be gathered.

**Panel 1.**
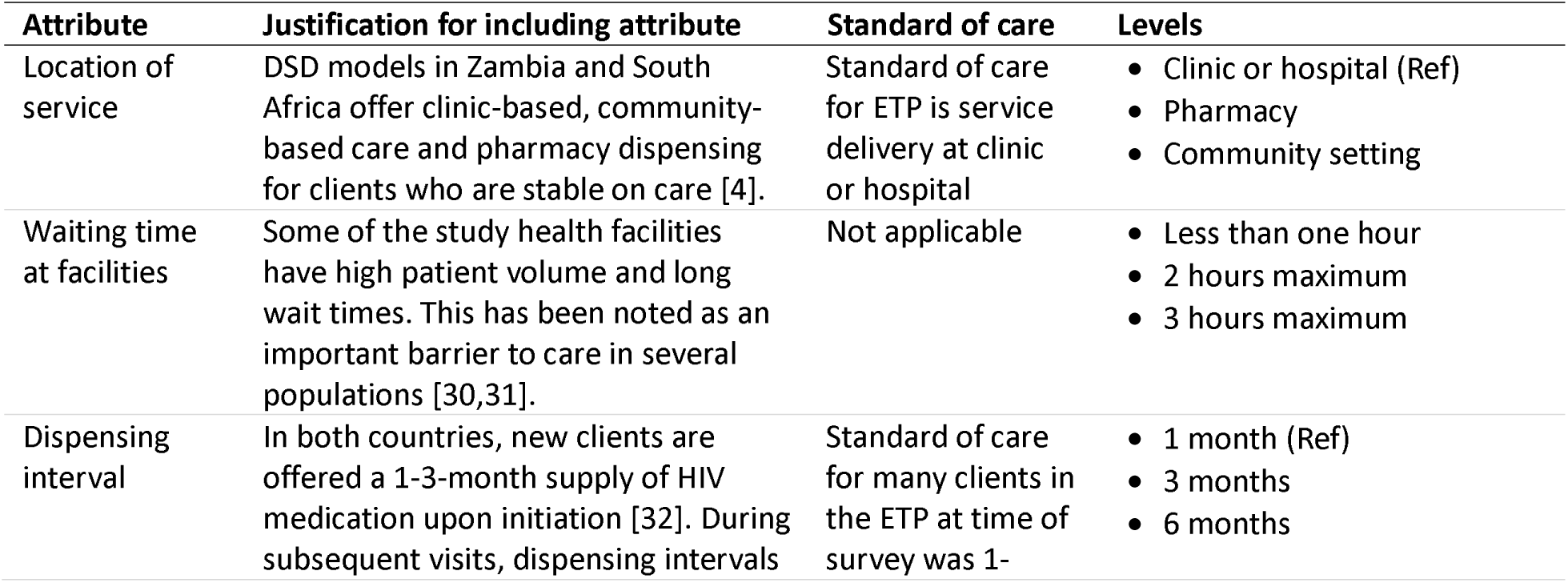
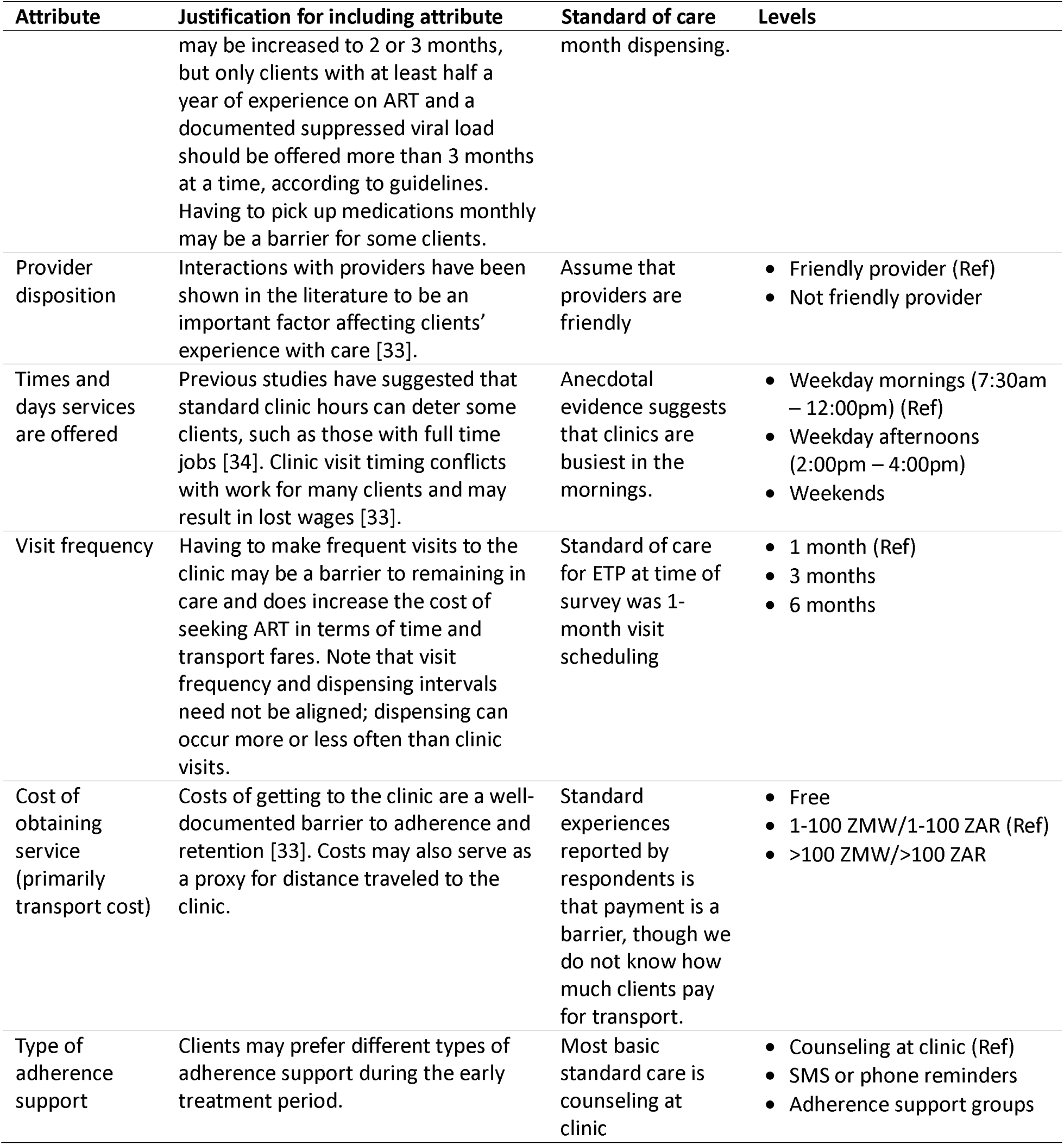
Attributes and levels.

| **Attribute** | **Justification for including attribute** | **Standard of care** | **Levels** |
| --- | --- | --- | --- |
| Location of service | DSD models in Zambia and South Africa offer clinic-based, community-based care and pharmacy dispensing for clients who are stable on care [4]. | Standard of care for ETP is service delivery at clinic or hospital | • Clinic or hospital (Ref)<br>• Pharmacy<br>• Community setting |
| Waiting time at facilities | Some of the study health facilities have high patient volume and long wait times. This has been noted as an important barrier to care in several populations [30,31]. | Not applicable | • Less than one hour<br>• 2 hours maximum<br>• 3 hours maximum |
| Dispensing interval | In both countries, new clients are offered a 1-3-month supply of HIV medication upon initiation [32]. During subsequent visits, dispensing intervals | Standard of care for many clients in the ETP at time of survey was 1- | • 1 month (Ref)<br>• 3 months<br>• 6 months |

| Attribute | Justification for including attribute | Standard of care | Levels |
| --- | --- | --- | --- |
|  | may be increased to 2 or 3 months, but only clients with at least half a year of experience on ART and a documented suppressed viral load should be offered more than 3 months at a time, according to guidelines. Having to pick up medications monthly may be a barrier for some clients. | month dispensing. |  |
| Provider disposition | Interactions with providers have been shown in the literature to be an important factor affecting clients' experience with care [33]. | Assume that providers are friendly | • Friendly provider (Ref)<br>• Not friendly provider |
| Times and days services are offered | Previous studies have suggested that standard clinic hours can deter some clients, such as those with full time jobs [34]. Clinic visit timing conflicts with work for many clients and may result in lost wages [33]. | Anecdotal evidence suggests that clinics are busiest in the mornings. | • Weekday mornings (7:30am – 12:00pm) (Ref)<br>• Weekday afternoons (2:00pm – 4:00pm)<br>• Weekends |
| Visit frequency | Having to make frequent visits to the clinic may be a barrier to remaining in care and does increase the cost of seeking ART in terms of time and transport fares. Note that visit frequency and dispensing intervals need not be aligned; dispensing can occur more or less often than clinic visits. | Standard of care for ETP at time of survey was 1-month visit scheduling | • 1 month (Ref)<br>• 3 months<br>• 6 months |
| Cost of obtaining service (primarily transport cost) | Costs of getting to the clinic are a well-documented barrier to adherence and retention [33]. Costs may also serve as a proxy for distance traveled to the clinic. | Standard experiences reported by respondents is that payment is a barrier, though we do not know how much clients pay for transport. | • Free<br>• 1-100 ZMW/1-100 ZAR (Ref)<br>• >100 ZMW/>100 ZAR |
| Type of adherence support | Clients may prefer different types of adherence support during the early treatment period. | Most basic standard care is counseling at clinic | • Counseling at clinic (Ref)<br>• SMS or phone reminders<br>• Adherence support groups |

For DCE analysis, we created dummy variables for each of the levels of all of the attributes. A reference level was selected for each attribute to represent the standard of care or commonly reported client experiences; the dummy variables for the reference levels were excluded from the model. Definitions of all variables are provided in Supplementary File 2. The DCE was analyzed using conditional logit model using STATA 17.0 (StataCorp 2023, Stata Statistical Software: Release 17, College Station, TX: StataCorp LLC.) [29]. Association between each attribute level and choice selection are reported as odds ratios with 95% confidence intervals.

### Focus group discussions and analysis

For the focus groups, interview guides were developed by the study investigators, informed by findings from the PREFER baseline survey. Guides addressed three main topics: i) the HIV treatment experience: did or did not go well during their early treatment experience, and why people start and stop treatment; ii) treatment preferences: what ideal service delivery would look like during the early treatment period and how they would prefer to collect medication and receive information about HIV and treatment; and iii) expectations of care: suggestions for clinics to make care easier for clients, advice for new clients, and facilitators and barriers to care seeking. Probes were used throughout the interview guide to elicit further responses.

The FGDs were run by two study team members (a note taker and facilitator), conducted in a local language, and audio-recorded. Audio recordings were translated into English and transcribed verbatim. Standard procedures for preserving confidentiality during the FGD and transcription were applied.

The qualitative codebook was created using an inductive-deductive approach [35]. Most codes were developed a priori using the FGD guide, and additional codes and child codes were added as necessary during the coding process to create a final analytic codebook. Two study team members double-coded an initial set of transcripts, reviewed them for consistency, and resolved discrepancies. The two team members then single-coded the remaining transcripts. Thematic analysis was conducted to understand patient treatment preference from a qualitative perspective. All coding and analysis were conducted in NVivo version 14. Results are presented in aggregate and using illustrative quotes, accompanied by participants’ demographic information.

### Data triangulation

We employed a convergent mixed-methods integration approach to triangulate data from the DCE and FGDs [36]. Quantitative and qualitative data were collected concurrently and analyzed independently using conditional logit modeling for the DCE and thematic analysis for the FGDs. Integration occurred at the interpretation stage, using a joint display [37] to identify areas of convergence, complementarity, and divergence. DCE results quantified the relative importance of service attributes, while FGDs provided contextual explanations and deeper insight into participants’ preferences and experiences.

### Ethical considerations

The PREFER study was approved by research ethics committees in Boston (USA), Johannesburg (South Africa), and Lusaka (Zambia): Boston University Institutional Review Board (H-42726, H-42903), University of the Witwatersrand Human Research Ethics Committee (M220440, M210342) and ERES-Converge IRB (2022-June-007, June 24, 2022). Regulatory approval was provided by Provincial Health Research Committees (National Health Research Database, South Africa) and the Zambia National Health Research Authority (NHRA000007/10/07/2022). All participants provided written informed consent. The study is registered with Clinicaltrials.gov as NCT05454839 (South Africa) and NCT05454852 (Zambia).

## Results

### Study sample characteristics

We enrolled 128 participants in Zambia (55% female, median age 35) and 122 in South Africa (SA) (83% female, median age 33) (Table 1). Supplementary Table 1 reports characteristics of the full PREFER sample alongside those enrolled in the DCE/FGD study activity who were selected from among those who reported or anticipated difficulties with adherence. Within the sample, formal employment was rare, with 60% of South Africans classifying themselves as unemployed and 52% of Zambians classifying themselves as informally employed. Most participants (63% in South Africa and 88% in Zambia) said that they would find it difficult to obtain the equivalent of $5-6 if needed to pay for healthcare.

**Table 1.**
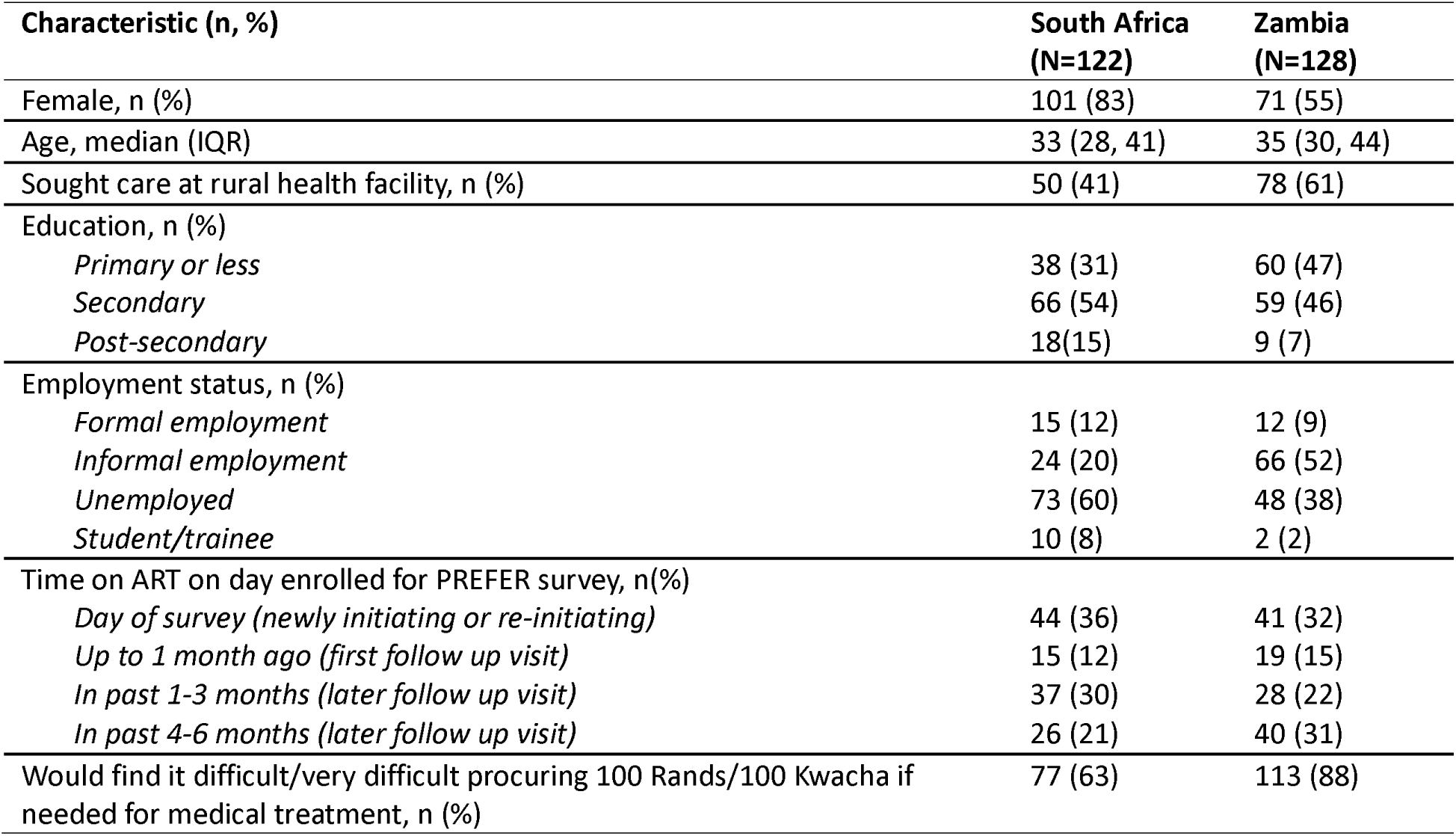
Respondent demographics.

| Characteristic (n, %) | South Africa<br>(N=122) | Zambia<br>(N=128) |
| --- | --- | --- |
| Female, n (%) | 101 (83) | 71 (55) |
| Age, median (IQR) | 33 (28, 41) | 35 (30, 44) |
| Sought care at rural health facility, n (%) | 50 (41) | 78 (61) |
| Education, n (%) |  |  |
| Primary or less | 38 (31) | 60 (47) |
| Secondary | 66 (54) | 59 (46) |
| Post-secondary | 18(15) | 9 (7) |
| Employment status, n (%) |  |  |
| Formal employment | 15 (12) | 12 (9) |
| Informal employment | 24 (20) | 66 (52) |
| Unemployed | 73 (60) | 48 (38) |
| Student/trainee | 10 (8) | 2 (2) |
| Time on ART on day enrolled for PREFER survey, n(%) |  |  |
| Day of survey (newly initiating or re-initiating) | 44 (36) | 41 (32) |
| Up to 1 month ago (first follow up visit) | 15 (12) | 19 (15) |
| In past 1-3 months (later follow up visit) | 37 (30) | 28 (22) |
| In past 4-6 months (later follow up visit) | 26 (21) | 40 (31) |
| Would find it difficult/very difficult procuring 100 Rands/100 Kwacha if needed for medical treatment, n (%) | 77 (63) | 113 (88) |

### DCE results

Preferences for attributes of HIV service delivery during the early treatment period are shown in Table 2. Respondents in both countries preferred receiving services in health facilities rather than through community-based care. Responses indicated a dose response to dispensing intervals: clients preference increased with increasing duration of dispensing intervals yet preferred more regular interactions with their provider during the first 6 months on treatment. Not unexpectedly, respondents, particularly in Zambia, were deterred from accessing services by unfriendly providers. Time spent waiting for services, time or day of week of services, and cost of getting to services did not influence choices in either country.

**Table 2.**
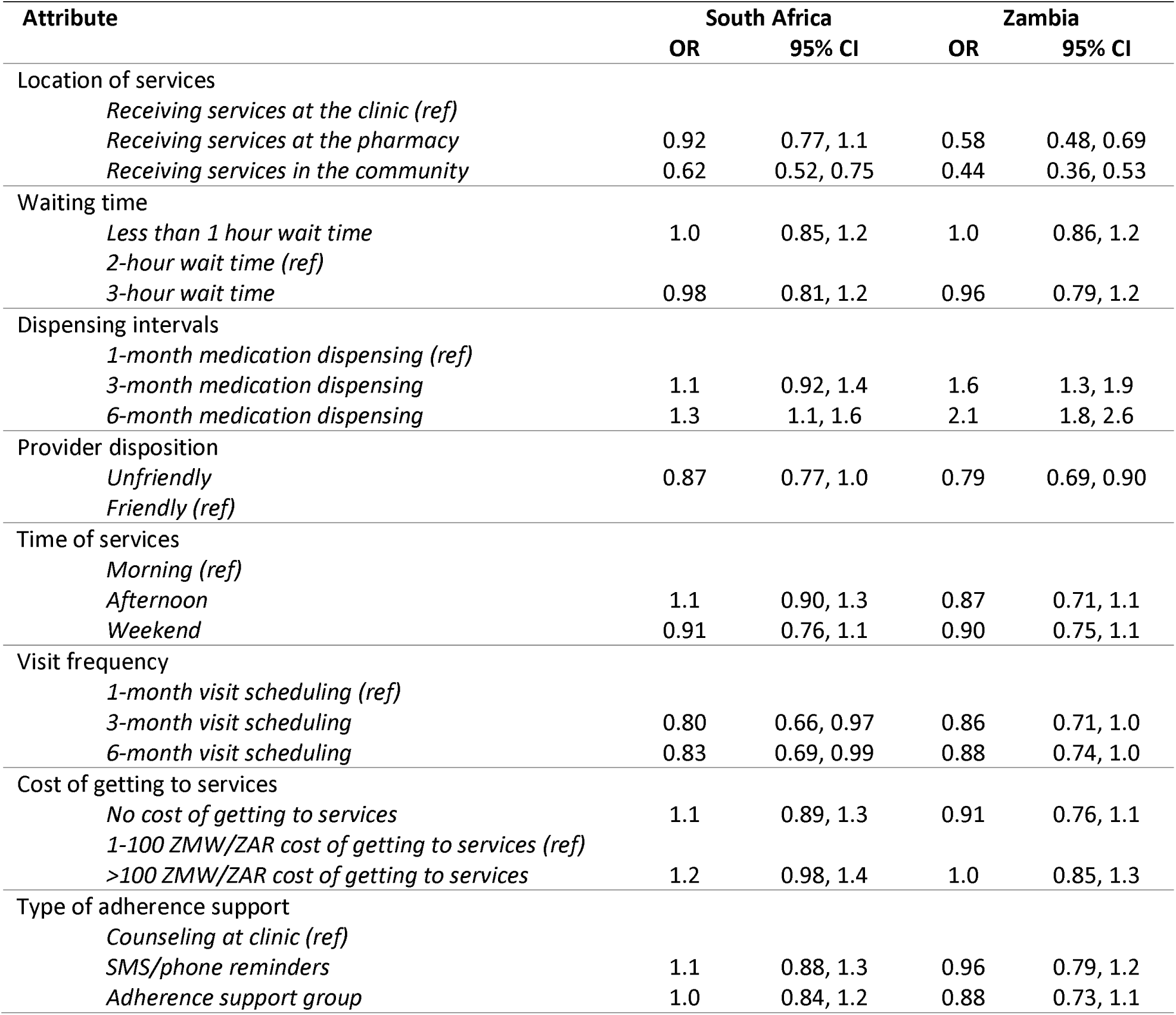
Odds ratios for client preferences for attributes of HIV service delivery during the early treatment period.

### Qualitative results

Consistent with the DCE findings in which respondents had strong preferences for clinic- or pharmacy-based services, respondents in both countries qualitatively described the benefits of facility-based services, including facility-based medication pickup. Participants in South Africa explained that new or re-initiating clients prefer to collect medication from the clinic, emphasizing the importance of the opportunity to ask questions and establish trust in the provider. Zambian participants noted that collecting at the clinic allows for closer medical attention and counseling.

> “When you are starting, the clinic is a preferable place. You will be able to ask and report things you are unsure of as you’ve started medication.” -Participant, South Africa

> “I would like to come for myself because I am still trying to get used to the idea that I am HIV positive. I also still have questions that I need to ask the nurses. I am also scared of the questions that I will get from the person who will pick for me. It is still sensitive.” -Participant, South Africa

> “If I go to the clinic there, I will have the health care provider attend to me properly and will also receive adequate counseling unlike some one just delivering for me at home.” -Participant, Zambia

Participants in Zambia acknowledged that while models of care exist that allow a trusted acquaintance to pick up medications on behalf of the participant, they generally preferred to pick up medications themselves. Participants expressed that picking up medication themselves fosters accountability and ownership and relieves them of relying on someone else for their care and adherence.

Qualitative findings revealed that long wait times and inefficient processes remained sources of frustration for participants accessing care at health facilities. Specifically, respondents in Zambia explained that misplaced files and time spent looking for them seemed inefficient, occupying facility staff time and attention. In South Africa, respondents described having to navigate a confusing clinic layout and to wait in several queues/rooms to receive care.

> “My challenge is that I used to take pills in room 1. Now they have moved me to take them to the other room. When I get there I am expected to join a queue even though I have already been in the queue.” Participant, South Africa

Participants had varying preferences surrounding medication dispensing in the first 6 months. Those in South Africa generally described positive opinions regarding 1–3-month dispensing intervals, while some in Zambia qualitatively expressed a preference for 6-month dispensing. Participants reported a wide variety of reasons to support their opinions.

> “The person who is resuming should do 1 month to 1 month. Just to test her.” – Participant, South Africa

> “I would be happy to receive three months’ supply from the pharmacy and only come [to clinic] to take bloods. This file issue is not pleasant, that is why people default.” – Participant, South Africa

> “3 months is normal because you are still facing a lot of challenges in your body. Sometimes you don’t have the strength. For you to manage…after 2 weeks to go back, it will be difficult for you.” – Participant, Zambia

> “I would prefer 6 months because it gives you some breathing space, you can even go to work without returning frequently at the clinic.” – Participant, Zambia

Participants in the FGDs articulated that they would prefer to be able to make clinic visits during non-working hours. Respondents in Zambia recommended that clinics extend service hours to include Saturdays, allowing patients to see the doctor and collect medications at a time with less clinic congestion. Respondents in South Africa reported a preference for medication collecting before or after work/school or on weekends.

Finally, participants in FGDs spoke to the importance of high quality, empathetic counselling, with a specific focus on provider disposition. FGD participants valued warm and empathetic interactions with providers, especially as it pertains to counselling. Respondents in Zambia were generally happy with the counseling they had received in the early treatment period, but reinforced the need for it to be frequent, empathetic, and clinic-based.

> “I think it is better if you get good counselling. To help you understand what is about to happen to you now that you will be taking medication…You need to find someone to motivate through the early days. Finding compassion at the clinic will help.” – Participant, South Africa

Respondents in Zambia also expressed a strong preference for peer support groups, explaining they used to be available and were valuable to people they knew.

### Data integration

Triangulating findings from the DCE and FGDs revealed both convergence and complementarity in client preferences for HIV service delivery during the early treatment period among a sample of those who reported having, or expected to have, challenges in the first six months. In both countries, DCE results showed a clear preference for receiving services at facilities rather than through community-based models. This was echoed in the FGDs, where participants explained that new or re-initiating clients value facility visits as opportunities to ask questions, get counselled, and build trust or rapport with their providers. This suggests the preference that emerged from the DCE is grounded in emotional safety and feeling supported, rather than convenience alone, illuminating why factors like time spent waiting for services, time or day of week of services, and cost of getting to services did not influence choices in either country in the DCE. While the DCE revealed an overall preference for longer dispensing intervals, FGD participants provided context for why this might not be preferred for all: some favored shorter intervals early in treatment for increased monitoring and support, while others appreciated the flexibility of six-month refills. Importantly, both methods confirmed the deterrent effect of unfriendly providers, with the DCE showing a strong negative response to poor provider disposition and FGDs underscoring the value of warm, empathetic care, particularly in the early stages of ART.

## Discussion

This study used a novel mixed-methods design, combining a DCE with concurrent FGDs to explore and explain HIV service delivery preferences during the early treatment period among clients in South Africa and Zambia. While DCEs are increasingly used in global health to quantify preferences and trade-offs, they are rarely paired with concurrent qualitative data collection using the same participants [9]. The concurrent design allowed us to interpret not only what clients preferred, but why, giving us a more enriched and nuanced understanding of client priorities during this critical window for engagement.

Across both countries in the DCE, clients expressed a clear preference for facility-based care during the early treatment period, findings that align with those from other studies among stable clients [22]. The strong preference for facility-based care was also reinforced in the FGDs, where participants emphasized the importance of asking questions, receiving personalized counseling, and building rapport with providers. These findings underscore the relational value of early facility visits that allow interaction with a higher cadre of health care worker, which is not always captured when considering interventions to improve efficiency which may task shift and move out of a facility. A similar phenomenon was also observed among stable clients not in the early treatment period in two DCEs in Kenya [17,22], and may reflect a broader perceived value of direct access to health care workers and a confidence or trust in the health system. While community-based ART delivery models remain a promising strategy for some populations [38–41], our findings suggest that early treatment models should retain intentional touchpoints with facilities and providers to meet clients’ needs for emotional support and information.

Our findings also highlight the complexity around preferences for dispensing intervals, which clients seem to have interpreted as separate from facility visits. This finding was also consistent with other DCEs conducted in the region, in which clients prefer longer dispensing intervals [21]. In both countries, while DCE results suggested a clear preference for longer dispensing intervals, FGD results were more varied, with some participants wanting closer monitoring and or had concerns about side effects. These stated preferences suggest convenience is driving the preference, but qualitative insights reveal concerns about readiness and side-effects suggesting that flexibility might be key to reach the needs of all. The 2023 South African National HIV Care and Treatment guidelines lowered the DSD eligibility threshold from six to four months [42], so clients in the early treatment period can have earlier access to longer-dispensing intervals offered in most models. Evaluation of the impact of these new guidelines, now underway, will provide empirical evidence against which to assess our participants’ self-reports.

Notably, positive provider interactions were consistently prioritized across both methods in both countries. In the DCE, unfriendly providers were a strong deterrent to care, while FGDs emphasized the importance of warmth, empathy, and respect in shaping the treatment experience. These results corroborate findings from prior studies showing that provider demeanor has a significant impact on patient satisfaction and retention [21,34]. However, improving provider attitudes remains challenging, given broader system constraints. Health care workers often operate in under-resourced, high-volume environments that may compromise their ability to provide empathetic care ; They report burnout, feeling overwhelmed and systemic barriers such as inadequate care delivery models and system capacity constraints [43,44]. Solutions intended to reduce provider burden—such as longer dispensing intervals that decrease patient volume or AI power health care assistants which support health care workers with triaging and administration—could improve both the quality of client-provider interactions and provider well-being.

Interestingly, operational factors like wait times, visit timing, and travel costs did not significantly drive preferences in the DCE in either South Africa or Zambia, despite being commonly cited in the FGDs as sources of frustration. Rather than indicating a conflict between methods, this divergence reflects how clients prioritize competing needs. In settings where unemployment is high and time pressures vary, waiting at a clinic may be irritating but not prohibitive, and may be deprioritized when weighed against relational and structural attributes like dispensing intervals or provider behavior. While the DCE captures decision-making across multiple attributes, offering a fuller picture of trade-offs that qualitative data alone may not predict, the FGDs help contextualize and richly describe preferences within individual attributes. In this case, findings are consistent with a robust qualitative literature that finds longer waiting time may decrease satisfaction with ART care [45], but the frustrations articulated do not ultimately drive their actual decision-making behavior. We also hypothesize that participants’ tolerance for long wait times may be linked to infrequent clinic visits. If a client only attends once every 3-6 months, the burden of wait time may feel more acceptable than if they attend monthly. This may explain why long lines and inefficient processes, while frustrating, were not expressed as drivers of decision-making when weighed against other factors.

Given the strong preference for facility-based care, longer dispensing intervals, and positive provider interactions across both countries, tailoring early treatment models to reflect these priorities appears not only feasible but potentially straightforward. In both South Africa and Zambia, clients expressed a desire to remain connected to facility-based services in early care – not because of convenience, but because of access to a trusted provider and high-quality counseling. The 2023 South African National HIV Care and Treatment guidelines included a revision to provide improved counselling at ART initiation to support engagement in care, underscoring the importance of high-quality support during this period [42]. These findings also suggest that shifting services into the community during this period may be unnecessary or even counterproductive, at least for the patients in this sample who had had, or anticipated having challenges in the early treatment period. Similarly, although clients preferred longer refill intervals even during the early months, this is primarily a logistical issue, not clinical, and may be addressed through supply chain changes. These changes could also reduce clinic volume, indirectly relieving pressure on providers who remain essential figures of the client’s clinical service experience but are often busy or overburdened. Our data suggest it may be prudent to prioritize access to skilled, compassionate providers at key touchpoints early in their treatment journey. Overall, client preferences illuminated here suggest that a more responsive early treatment model could be created by adapting existing facility-based care, extending refill options, and prioritizing provider quality[46] as a core component of client-centered care.

## Conclusion

Despite the limitations noted above, our study’s approach of combining quantitative preference data with qualitative narratives from the same participants allowed us to gain a more nuanced understanding of preferences and how clients prioritize these preferences than either method would have supported alone. Our findings support the case for more flexible, client-centered approaches to ART delivery in the early treatment period. To date, most DCE research on DSD models has focused on stable clients, with limited exploration of preferences during the early treatment period. As countries like South Africa adopt revised guidelines that expand DSD eligibility to clients earlier in treatment, it is increasingly important to understand whether the models developed for stable clients align with the needs of those newly initiating or re-initiating ART. Our findings suggest that the early treatment period is experienced differently—marked by a need for support, trust-building, and reassurance—yet clients also express readiness for convenience-enhancing features like longer dispensing intervals. Tailoring DSD models to this early phase requires balancing relational support with structural flexibility. By identifying which components of DSD are both acceptable and supportive from the outset, programs can better align with client preferences and may reduce early disengagement from care.

## Supporting information

Supplementary File 1. Choice sets

Supplementary File 2. Variable definitions

Supplementary Table 1. Full PREFER demographic characteristics

## Authors’ contributions

## Acknowledgements

We would like to thank the clients and staff of the study sites for their cooperation in allowing us to conduct this study and the South Africa National Department of Health and Zambian Ministry of Health for approving this research.

## Data availability

Data generated by the study will be made publicly available in the Open BU repository (https://open.bu.edu/) after the PREFER study protocol has been closed (anticipated closure December 2026). Until then, data will remain under the supervision of the Boston University Medical Campus IRB, the University of the Witwatersrand Human Research Ethics Committee (HREC), and the ERES-Converge IRB. Requests can be sent to the BUMC IRB at.

## Ethics

The PREFER study was approved by research ethics committees in Boston (USA), Johannesburg (South Africa), and Lusaka (Zambia): Boston University Institutional Review Board (H-42726, H-42903), University of the Witwatersrand Human Research Ethics Committee (M220440, M210342) and ERES-Converge IRB (2022-June-007, June 24, 2022). Regulatory approval was provided by Provincial Health Research Committees (National Health Research Database, South Africa) and the Zambia National Health Research Authority (NHRA000007/10/07/2022). All participants provided written informed consent. The study is registered under Clinicaltrials.gov as NCT05454839 (South Africa) and NCT05454852 (Zambia).

## Supplementary Materials

Supplementary File 1. All choice sets

Supplementary File 2. Variable definitions

**Supplementary Table 1.** Demographic and socio-economic characteristics of enrolled participants.

| Characteristic (n, %) | South Africa<br>PREFER full sample<br>(N=1,098) | Qualitative<br>FGD sample<br>South Africa<br>(N=122) | Zambia<br>PREFER full<br>sample<br>(N=771) | Zambia<br>(N=128) |
| --- | --- | --- | --- | --- |
| Female, n (%) | 786 (72) | 101 (83) | 514 (67) | 71 (55) |
| Age, median (IQR) | 33 (27, 41) | 33 (28, 41) | 32 (27, 40) | 35 (30, 44) |
| Sought care at rural health facility, n (%) | 506(46) | 50 (41) | 623 (81) | 78 (61) |
| Education, n (%) |  |  |  |  |
| Primary or less | 409 (37) | 38 (31) | 386 (50) | 60 (47) |
| Secondary | 525 (48) | 66 (54) | 329 (43) | 59 (46) |
| Post-secondary | 164 (15) | 18(15) | 56 (7) | 9 (7) |
| Employment status, n (%) |  |  |  |  |
| Formal employment | 240 (22) | 15 (12) | 73 (9) | 12 (9) |
| Informal employment | 216 (20) | 24 (20) | 413 (54) | 66 (52) |
| Unemployed | 562 (51) | 73 (60) | 267 (35) | 48 (38) |
| Student/trainee | 80 (7) | 10 (8) | 18 (2) | 2 (2) |
| Time on ART on day enrolled for<br>PREFER baseline survey, n(%) |  |  |  |  |
| Day of survey (newly initiating or re-initiating) | 419 (38) | 44 (36) | 261 (34) | 41 (32) |
| Up to 1 month ago (first follow up<br>visit) | 170 (15) | 15 (12) | 115 (15) | 19 (15) |
| In past 1-3 months (later follow up<br>visit) | 249 (23) | 37 (30) | 172 (22) | 28 (22) |
| In past 4-6 months (later follow up<br>visit) | 260 (24) | 26 (21) | 223 (29) | 40 (31) |
| Would find it difficult/very difficult<br>procuring 100 Rands if needed for<br>medical treatment, n (%) | 615 (56) | 77 (63) | 629 (81) | 113 (88) |

## References

1. World Health Organization. The final stretch to reaching 95-95-95 targets: supporting re-engagement in HIV treatment. 22 Jul 2024 [cited 28 Aug 2024]. Available: https://www.who.int/news/item/22-07-2024-the-final-stretch-to-reaching-95-95-95-targets--supporting-re-engagement-in-hiv-treatment

2. Rosen S, Grimsrud A, Ehrenkranz P, Katz I. Models of service delivery for optimizing a patient’s first six months on antiretroviral therapy for HIV: an applied research agenda. Gates Open Res. 2020;4: 1–15. doi:10.12688/GATESOPENRES.13159.1

3. Burke RM, Rickman HM, Pinto C, Ehrenkranz P, Choko A, Ford N. Reasons for disengagement from antiretroviral care in the era of “Treat All” in low-or middle-income countries: a systematic review. J Int AIDS Soc. 2024;27: 26230. doi:10.1002/jia2.26230/full

4. Huber A, Pascoe S, Nichols B, Long L, Kuchukhidze S, Phiri B, et al. Differentiated Service Delivery Models for HIV Treatment in Malawi, South Africa, and Zambia: A Landscape Analysis. Glob Health Sci Pract. 2021;9: 296–307. doi:10.9745/GHSP-D-20-00532

5. De Bekker-Grob EW, Ryan M, Gerard K. Discrete choice experiments in health economics: a review of the literature. Health Econ. 2012;21: 145–172. doi:10.1002/HEC.1697

6. Lancsar E, Louviere J. Conducting discrete choice experiments to inform healthcare decision making: A user’s guide. Pharmacoeconomics. 2008;26: 661–677. doi:10.2165/00019053-200826080-00004,

7. Merton RK, Kendall PL. The Focused Interview. https://doi.org/101086/219886. 1946;51: 541–557. doi:10.1086/219886

8. Green J, Thorogood N, editors. Qualitative Methods for Health Research. 4th ed. SAGE Publications Ltd; 2014.

9. Vass C, Rigby D, Payne K. The Role of Qualitative Research Methods in Discrete Choice Experiments: A Systematic Review and Survey of Authors. [cited 24 Feb 2025]. doi:10.1177/0272989X16683934

10. Coast J, Al-Janabi H, Sutton EJ, Horrocks SA, Vosper AJ, Swancutt DR, et al. Using qualitative methods for attribute development for discrete choice experiments: Issues and recommendations. Health Econ. 2012;21: 730–741. doi:10.1002/HEC.1739,

11. Mangham LJ, Hanson K, McPake B. How to do (or not to do) … Designing a discrete choice experiment for application in a low-income country. Health Policy Plan. 2009;24: 151–158. doi:10.1093/HEAPOL/CZN047

12. Kløjgaard ME, Bech M, Søgaard R. Designing a Stated Choice Experiment: The Value of a Qualitative Process. Journal of Choice Modelling. 2012;5: 1–18. doi:10.1016/S1755-5345(13)70050-2

13. Govathson C, Long L, Moolla A, Mngadi-Ncube S, Ngcobo N, Mongwenyana C, et al. Understanding school-going adolescent’s preferences for accessing HIV and contraceptive care: findings from a discrete choice experiment among learners in Gauteng, South Africa. BMC Health Serv Res. 2023;23: 1378. doi:10.1186/S12913-023-10414-W

14. Youssef E, Cooper V, Miners A, Llewellyn C, Pollard A, Lagarde M, et al. Understanding HIV-positive patients’ preferences for healthcare services: a protocol for a discrete choice experiment. BMJ Open. 2016;6. doi:10.1136/BMJOPEN-2015-008549

15. Ong JJ, Nwaozuru U, Obiezu-Umeh C, Airhihenbuwa C, Xian H, Terris-Prestholt F, et al. Designing HIV testing and self-testing services for young people in Nigeria: A discrete choice experiment. Patient. 2021;14: 815. doi:10.1007/S40271-021-00522-2

16. Chiwire P, Mühlbacher AC, Evers SM, Mahomed H, Ostermann J, Hiligsmann M. A discrete choice experiment investigating HIV testing preferences in South Africa. J Med Econ. 2022;25: 481–490. doi:10.1080/13696998.2022.2055937

17. Mando RO, Moghadassi M, Juma E, Ogollah C, Packel L, Kulzer JL, et al. Patient preferences for HIV service delivery models; a Discrete Choice Experiment in Kisumu, Kenya. PLOS Global Public Health. 2022;2: e0000614. doi:10.1371/JOURNAL.PGPH.0000614

18. Eshun-wilson I, Mukumbwa-mwenechanya M, Kim H, Zanolini AL, Mwamba C, Dowdy D, et al. Differentiated Care Preferences of Stable Patients on ART in Zambia. CROI 2019. 2019;Abstract 1: 1054.

19. Mugambi ML, Odhiambo BO, Dollah A, Marwa MM, Nyakina J, Kinuthia J, et al. Women’s preferences for HIV prevention service delivery in pharmacies during pregnancy in Western Kenya: a discrete choice experiment. J Int AIDS Soc. 2024;27. doi:10.1002/JIA2.26301/FULL

20. Goswami S, Bentley JP, Kang M, Bhattacharya K, Barnard M. Preferences for a community pharmacy-based pre-exposure prophylaxis (PrEP) delivery program: A discrete choice experiment. Journal of the American Pharmacists Association. 2024;64. doi:10.1016/j.japh.2024.102091

21. Zanolini A, Sikombe K, Sikazwe I, Eshun-Wilson I, Somwe P, Bolton Moore C, et al. Understanding preferences for HIV care and treatment in Zambia: Evidence from a discrete choice experiment among patients who have been lost to follow-up. PLoS Med. 2018;15: e1002636. doi:10.1371/JOURNAL.PMED.1002636

22. Dommaraju S, Hagey J, Odeny TA, Okaka S, Kadima J, Bukusi EA, et al. Preferences of people living with HIV for differentiated care models in Kenya: A discrete choice experiment. PLoS One. 2021;16: e0255650. doi:10.1371/JOURNAL.PONE.0255650

23. Maskew M, Ntjikelane V, Juntunen A, Scott N, Benade M, Sande L, et al. Preferences for services in a patient’s first six months on antiretroviral therapy for HIV in South Africa and Zambia (PREFER): research protocol for a prospective observational cohort study. Gates Open Res. 2024;7: 119. doi:10.12688/GATESOPENRES.14682.2

24. Mutanda N, Morgan A, Kamanga A, Sande L, Ntjikelane V, Maskew M, et al. Experiences and Preferences in Zambia and South Africa for Delivery of HIV Treatment During a Client’s First Six Months: Results of the PREFER Study’s Cross-Sectional Baseline Survey. AIDS Behav. 2025 [cited 18 Feb 2025]. doi:10.1007/S10461-025-04640-Y

25. Hennink MM, Kaiser BN, Weber MB. What Influences Saturation? Estimating Sample Sizes in Focus Group Research. Qual Health Res. 2019;29: 1483. doi:10.1177/1049732318821692

26. Speckemeier C, Krabbe L, Schwenke S, Wasem J, Buchberger B, Neusser S. Discrete choice experiment to determine preferences of decision-makers in healthcare for different formats of rapid reviews. Syst Rev. 2021;10: 1–8. doi:10.1186/S13643-021-01647-Z/TABLES/4

27. Friedel JE, Foreman AM, Wirth O. An introduction to “discrete choice experiments” for behavior analysts. Behavioural Processes. 2022;198: 104628. doi:10.1016/J.BEPROC.2022.104628

28. Ride J, Goranitis I, Meng Y, LaBond C, Lancsar E. A Reporting Checklist for Discrete Choice Experiments in Health: The DIRECT Checklist. Pharmacoeconomics. 2024;42: 1161. doi:10.1007/S40273-024-01431-6

29. Ryan M, Kolstad JR, Rockers PC, Dolea C. How to Conduct a Discrete Choice Experiment for Health Workforce Recruitment and Retention in Remote and Rural Areas: A User Guide With Case Studies (English). Washington, DC; 2012 Dec. Available: http://documents.worldbank.org/curated/en/586321468156869931

30. Ogbonnaya IN, Reed E, Wanyenze RK, Wagman JA, Silverman JG, Kiene SM. Perceived Barriers to HIV Care and Viral Suppression Comparing Newly Diagnosed Women Living with HIV in Rural Uganda with and without a History of Intimate Partner Violence. J Interpers Violence. 2022;37: NP17133–NP17156. doi:10.1177/08862605211028284

31. Ogunbajo A, Kershaw T, Kushwaha S, Boakye F, Wallace-Atiapah ND, Nelson LRE. Barriers, Motivators, and Facilitators to Engagement in HIV Care Among HIV-infected Ghanaian Men who have Sex with Men (MSM). AIDS Behav. 2018;22: 829. doi:10.1007/S10461-017-1806-6

32. Rosen S. The First Six Months of HIV Treatment: Clinic Visit Schedule and Dispensing Intervals at Zambian Clinics. 2022. Available: https://sites.bu.edu/ambit/

33. Sikazwe I, Eshun-Wilson I, Sikombe K, Beres LK, Somwe P, Mody A, et al. Patient-reported Reasons for Stopping Care or Switching Clinics in Zambia: A Multisite, Regionally Representative Estimate Using a Multistage Sampling-based Approach in Zambia. Clin Infect Dis. 2021;73: E2294–E2302. doi:10.1093/CID/CIAA1501

34. Bogart LM, Chetty S, Giddy J, Sypek A, Sticklor L, Walensky RP, et al. Barriers to care among people living with HIV in South Africa: contrasts between patient and healthcare provider perspectives. AIDS Care. 2013;25: 843–853. doi:10.1080/09540121.2012.729808

35. Proudfoot K. Inductive/Deductive Hybrid Thematic Analysis in Mixed Methods Research. J Mix Methods Res. 2023;17: 308–326. doi:10.1177/15586898221126816/ASSET/23B6B5FA-FA38-49E9-8F3A-47954BB112EC/ASSETS/IMAGES/10.1177_15586898221126816-IMG1.PNG

36. Erzberger C, Kelle U. Making Inferences in Mixed Methods: The Rules of Integration. In: Tashakkori A, Teddlie C, editors. Handbook of Mixed Methods in Social & Behavioral Research. SAGE Publications; 2003. Available: https://books.google.com/books?hl=en&lr=&id=F8BFOM8DCKoC&oi=fnd&pg=PA457&ots=gXcSuFowJd&sig=Jt2Cv_gn-Dxp9MyqUajy-OM_VQs#v=onepage&q&f=false

37. Guetterman TC, Fetters MD, Creswell JW. Integrating Quantitative and Qualitative Results in Health Science Mixed Methods Research Through Joint Displays. The Annals of Family Medicine. 2015;13: 554–561. doi:10.1370/AFM.1865

38. Ibiloye O, Masquillier C, Jwanle P, Van Belle S, van Olmen J, Lynen L, et al. Community-Based ART Service Delivery for Key Populations in Sub-Saharan Africa: Scoping Review of Outcomes Along the Continuum of HIV Care. AIDS Behav. 2022;26: 2314–2337. doi:10.1007/S10461-021-03568-3/TABLES/9

39. Barnabas R V., Szpiro AA, van Rooyen H, Asiimwe S, Pillay D, Ware NC, et al. Community-based antiretroviral therapy versus standard clinic-based services for HIV in South Africa and Uganda (DO ART): a randomised trial. Lancet Glob Health. 2020;8: e1305–e1315. doi:10.1016/S2214-109X(20)30313-2

40. Lippman SA, Pettifor A, Dufour MSK, Kabudula CW, Twine R, Peacock D, et al. A community mobilisation intervention to improve engagement in HIV testing, linkage to care, and retention in care in South Africa: a cluster-randomised controlled trial. Lancet HIV. 2022;9: e617–e626. doi:10.1016/S2352-3018(22)00192-8

41. Lifson AR, Hailemichael A, Workneh S, MacLehose RF, Horvath KJ, Hilk R, et al. A three-year randomized community trial of community support workers in rural Ethiopia to promote retention in HIV care. AIDS Care. 2022;34: 1506–1512. doi:10.1080/09540121.2022.2029819

42. Republic of South Africa National Department of Health. 2023 ART Clinical Guidelines for the Management of HIV in Adults, Pregnancy and Breastfeeding, Adolescents, Children, Infants and Neonates. 2023. Available: https://knowledgehub.health.gov.za/system/files/elibdownloads/2023-07/National%20ART%20Clinical%20Guideline%20AR%204.5%2020230713%20Version%204%20WEB.pdf

43. Kruse GR, Chapula BT, Ikeda S, Nkhoma M, Quiterio N, Pankratz D, et al. Burnout and use of HIV services among health care workers in Lusaka District, Zambia: A cross-sectional study. Hum Resour Health. 2009;7. doi:10.1186/1478-4491-7-55,

44. Atukunda R, Memiah P, Shumba C. Care for the caregiver: Stress relief and burnout among health workers in HIV care. Global Journal of Medicine and Public Health. 2013;2. Available: https://www.researchgate.net/publication/256427086_Care_for_the_caregiver_Stress_relief_and_burnout_among_health_workers_in_HIV_care

45. Nwagbara UI, Hlongwana KW, Chima SC. Mapping evidence on the factors contributing to long waiting times and interventions to reduce waiting times within primary health care facilities in South Africa: A scoping review. PLoS One. 2024;19: e0299253. doi:10.1371/JOURNAL.PONE.0299253

46. Sikombe K, Mody A, Goss CW, Simbeza S, Beres LK, Pry JM, et al. Effect of a multicomponent, person-centred care intervention on client experience and HIV treatment outcomes in Zambia: a stepped-wedge, cluster-randomised trial. Lancet HIV. 2025;12: e26–e39. doi:10.1016/S2352-3018(24)00264-9

