## Supplementary File 1. Choice sets for "Client preferences for service delivery during the early treatment period in South Africa and Zambia: Mixed-method findings from a discrete choice experiment and concurrent focus group discussions"

*South Africa DCE*
*Block 1*

### Choice Set 1

Block 1

#### Which option would you choose?

| Characteristic | Option A | Option B |
|---|---|---|
| Location of Service | Clinic/hospital image<br>Clinic or hospital | Community image<br>Community setting |
| The waiting time to get ART | 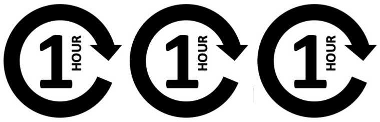<br>3 hour maximum | 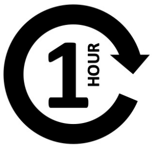<br>Less than 1 hour |
| The number of months of ART dispensed at a given time | 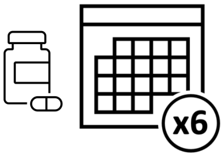<br>6 months of ART | 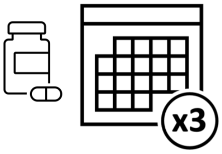<br>3 months of ART |
| Whether the provider is friendly or not | Provider image<br>Not friendly provider | Provider image<br>Friendly provider |
| The time of day you can get services | 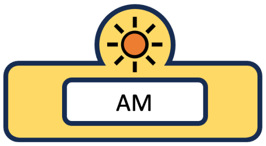<br>Weekday morning (7:30am to 12pm) | 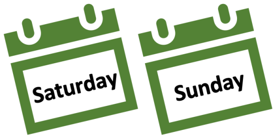<br>Weekends |
| How often visits are required (visit frequency) | 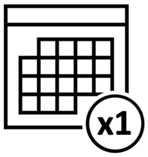<br>1 month | 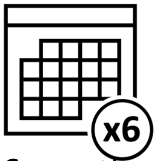<br>6 months |
| The cost of getting to the service | 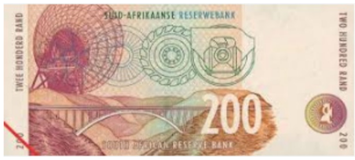<br>100-200 ZAR | 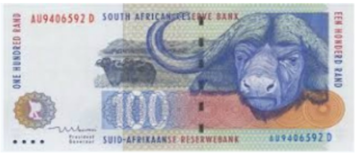<br>1-100 ZAR |
| Type of adherence support | 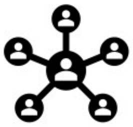<br>Adherence support groups | 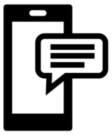<br>SMS/phone reminders |

### Choice Set 2

Block 1

#### Which option would you choose?

| Characteristic | Option A | Option B |
| --- | --- | --- |
| Location of Service | Pharmacy image<br>Pharmacy | Community image<br>Clinic or hospital |
| The waiting time to get ART | Less than 1 hour | 3 hour maximum |
| The number of months of ART dispensed at a given time | 3 months of ART | 6 months of ART |
| Whether the provider is friendly or not | Provider image<br>Not friendly provider | Provider image<br>Friendly provider |
| The time of day you can get services | PM<br>Weekday afternoon (2pm to 4pm) | AM<br>Weekday morning (7:30am to 12pm) |
| How often visits are required (visit frequency) | 6 months | 3 months |
| The cost of getting to the service | 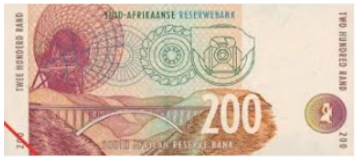<br>100-200 ZAR | 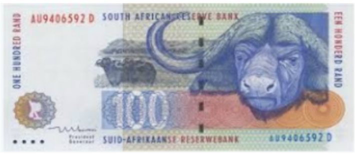<br>1-100 ZAR |
| Type of adherence support | Adherence support groups | Counseling at clinic |

### Choice Set 3

Block 1

#### Which option would you choose?

| Characteristic | Option A | Option B |
|---|---|---|
| Location of Service | Pharmacy image<br>Community setting | Community image<br>Pharmacy |
| The waiting time to get ART | 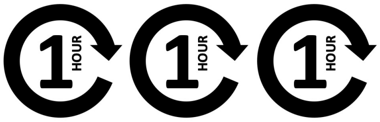 <br>3 hour maximum | 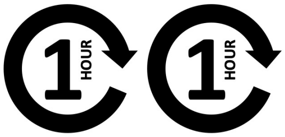 <br>2 hour maximum |
| The number of months of ART dispensed at a given time | 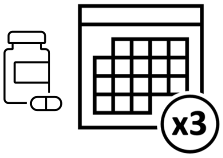 <br>3 months of ART | 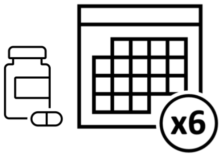 <br>6 months of ART |
| Whether the provider is friendly or not | Provider image<br>Friendly provider | Provider image<br>Not friendly provider |
| The time of day you can get services | 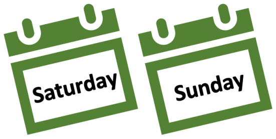 <br>Weekends | 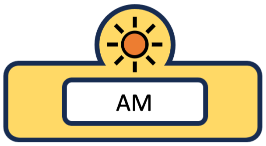 <br>Weekday morning (7:30am to 12pm) |
| How often visits are required (visit frequency) | 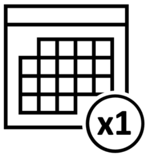 <br>1 month | 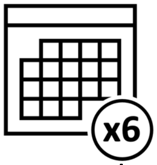 <br>6 months |
| The cost of getting to the service | 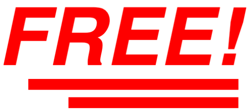 <br>No cost | 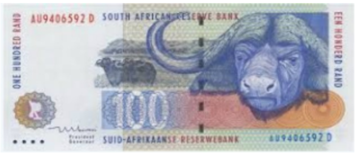<br>1-100 ZAR |
| Type of adherence support | 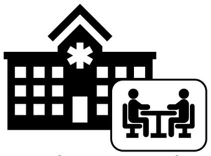<br>Counseling at clinic | 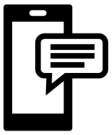<br>SMS/phone reminders |

*South Africa DCE*
*Block 1*

### <u>Choice Set 4</u>

Block 1

#### Which option would you choose?

| Characteristic | Option A | Option B |
|---|---|---|
| Location of Service | Pharmacy image<br>Clinic or hospital | Community image<br>Community setting |
| The waiting time to get ART | 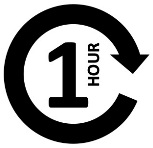<br>Less than 1 hour | 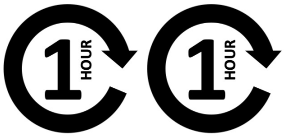<br>2 hour maximum |
| The number of months of ART dispensed at a given time | 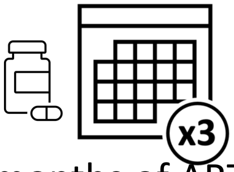<br>3 months of ART | 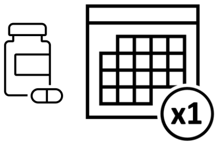<br>1 month of ART |
| Whether the provider is friendly or not | Provider image<br>Friendly provider | Provider image<br>Not friendly provider |
| The time of day you can get services | 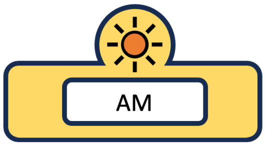<br>Weekday morning (7:30am to 12pm) | 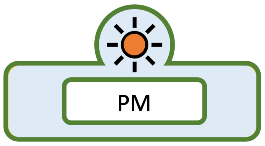<br>Weekday afternoon (2pm to 4pm) |
| How often visits are required (visit frequency) | 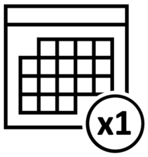<br>1 month | 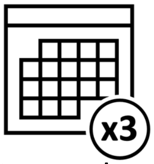<br>3 months |
| The cost of getting to the service | 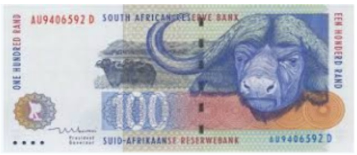<br>1-100 ZAR | 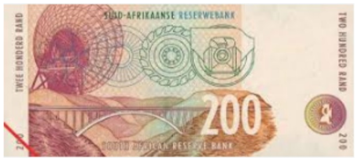<br>100-200 ZAR |
| Type of adherence support | 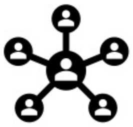<br>Adherence support groups | 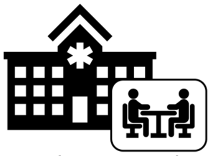<br>Counseling at clinic |

*South Africa DCE*
*Block 1*

### Choice Set 5

Block 1

#### Which option would you choose?

| Characteristic | Option A | Option B |
|---|---|---|
| Location of Service | Pharmacy image<br>Pharmacy | Community image<br>Clinic or hospital |
| The waiting time to get ART | 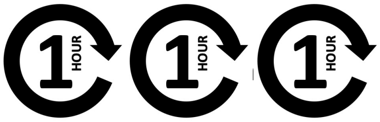<br>3 hour maximum | 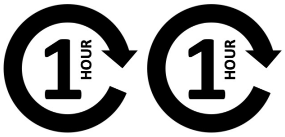<br>2 hour maximum |
| The number of months of ART dispensed at a given time | 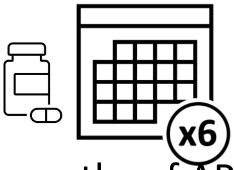<br>6 months of ART | 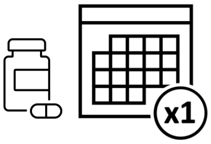<br>1 month of ART |
| Whether the provider is friendly or not | Provider image<br>Friendly provider | Provider image<br>Not friendly provider |
| The time of day you can get services | 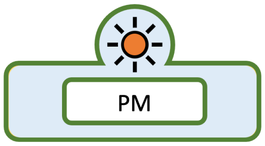<br>Weekday afternoon (2pm to 4pm) | 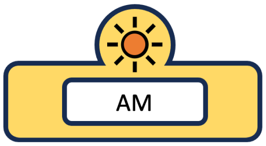<br>Weekday morning (7:30am to 12pm) |
| How often visits are required (visit frequency) | 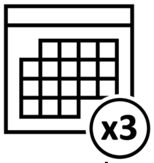<br>3 months | 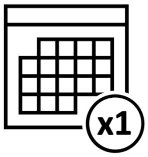<br>1 month |
| The cost of getting to the service | 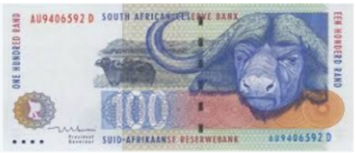<br>1-100 ZAR | **FREE!**<br>No cost |
| Type of adherence support | 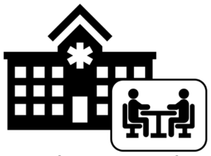<br>Counseling at clinic | 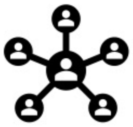<br>Adherence support groups |

### Choice Set 6

Block 1

Which option would you choose?

| Characteristic | Option A | Option B |
| --- | --- | --- |
| Location of Service | Pharmacy image<br>Pharmacy | Community image<br>Clinic or hospital |
| The waiting time to get ART | 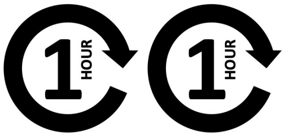<br>2 hour maximum | 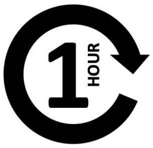<br>Less than 1 hour |
| The number of months of ART dispensed at a given time | 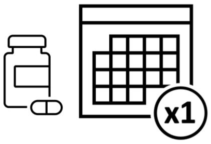<br>1 month of ART | 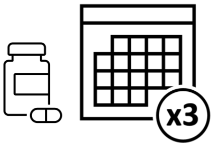<br>3 months of ART |
| Whether the provider is friendly or not | Provider image<br>Not friendly provider | Provider image<br>Friendly provider |
| The time of day you can get services | 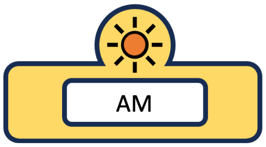<br>Weekday morning (7:30am to 12pm) | 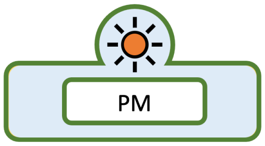<br>Weekday afternoon (2pm to 4pm) |
| How often visits are required (visit frequency) | 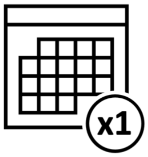<br>1 month | 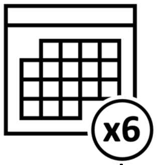<br>6 months |
| The cost of getting to the service | FREE!<br>No cost | 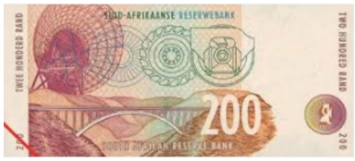<br>100-200 ZAR |
| Type of adherence support | 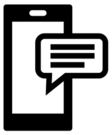<br>SMS/phone reminders | 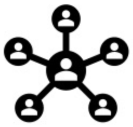<br>Adherence support groups |

### Choice Set 7

Block 1

#### Which option would you choose?

| Characteristic | Option A | Option B |
|---|---|---|
| Location of Service | Pharmacy image<br>Clinic or hospital | Community image<br>Community setting |
| The waiting time to get ART | 1 HOUR 1 HOUR<br>2 hour maximum | 1 HOUR 1 HOUR 1 HOUR<br>3 hour maximum |
| The number of months of ART dispensed at a given time | x3<br>3 months of ART | x6<br>6 months of ART |
| Whether the provider is friendly or not | Provider image<br>Friendly provider | Provider image<br>Not friendly provider |
| The time of day you can get services | PM<br>Weekday afternoon (2pm to 4pm) | Saturday Sunday<br>Weekends |
| How often visits are required (visit frequency) | x3<br>3 months | x6<br>6 months |
| The cost of getting to the service | 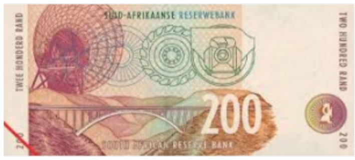<br>100-200 ZAR | FREE!<br>No cost |
| Type of adherence support | SMS/phone reminders | Adherence support groups |

### Choice Set 8

Block 1

#### Which option would you choose?

| Characteristic | Option A | Option B |
|---|---|---|
| Location of Service | Pharmacy image<br>Community setting | Community image<br>Clinic or hospital |
| The waiting time to get ART | 1 HOUR 1 HOUR<br>2 hour maximum | 1 HOUR<br>Less than 1 hour |
| The number of months of ART dispensed at a given time | x6<br>6 months of ART | x1<br>1 month of ART |
| Whether the provider is friendly or not | Provider image<br>Friendly provider | Provider image<br>Not friendly provider |
| The time of day you can get services | AM<br>Weekday morning (7:30am to 12pm) | Saturday Sunday<br>Weekends |
| How often visits are required (visit frequency) | x6<br>6 months | x3<br>3 months |
| The cost of getting to the service | FREE!<br>No cost | 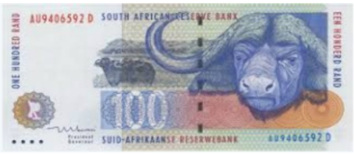<br>1-100 ZAR |
| Type of adherence support | SMS/phone reminders | Counseling at clinic |

### Choice Set 9



Which option would you choose?

| Characteristic | Option A | Option B |
|---|---|---|
| Location of Service | Pharmacy image<br>Pharmacy | Community image<br>Community setting |
| The waiting time to get ART | 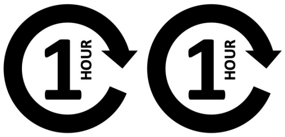 <br>2 hour maximum | 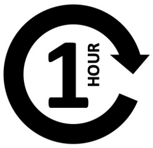 <br>Less than 1 hour |
| The number of months of ART dispensed at a given time | 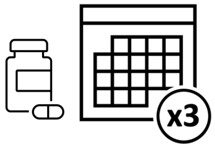 <br>3 months of ART | 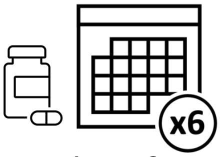 <br>6 months of ART |
| Whether the provider is friendly or not | Provider image<br>Not friendly provider | Provider image<br>Friendly provider |
| The time of day you can get services | 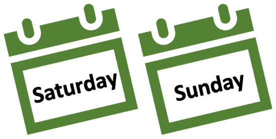 <br>Weekends | 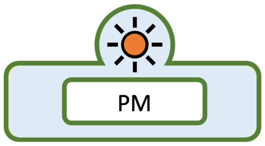 <br>Weekday afternoon (2pm to 4pm) |
| How often visits are required (visit frequency) | 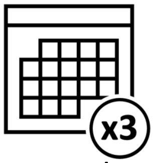 <br>3 months | 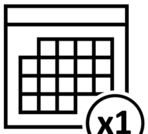 <br>1 month |
| The cost of getting to the service | 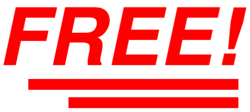 <br>No cost | 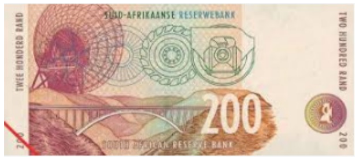 <br>100-200 ZAR |
| Type of adherence support | 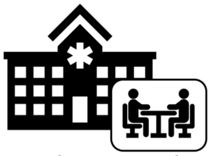<br>Counseling at clinic | 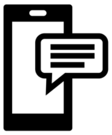<br>SMS/phone reminders |

### Choice Set 1

Block 2

Which option would you choose?

| Characteristic | Option A | Option B |
|---|---|---|
| Location of Service | Pharmacy image<br>Clinic or hospital | Community image<br>Pharmacy |
| The waiting time to get ART | 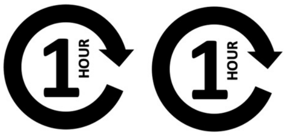<br>2 hour maximum | 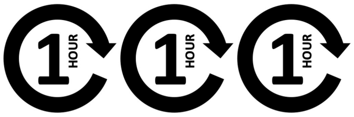<br>3 hour maximum |
| The number of months of ART dispensed at a given time | 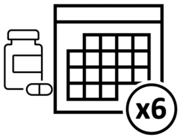 <br>6 month | 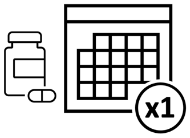 <br>1 month |
| Whether the provider is friendly or not | Provider image<br>Not friendly provider | Provider image<br>Friendly provider |
| The time of day you can get services | 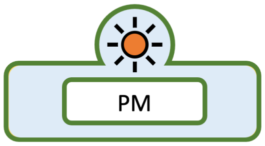 <br>Weekday afternoon (2pm to 4pm) | 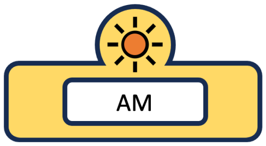 <br>Weekday morning (7:30am to 12pm) |
| How often visits are required (visit frequency) | 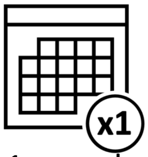 <br>1 month | 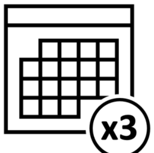 <br>3 months |
| The cost of getting to the service | 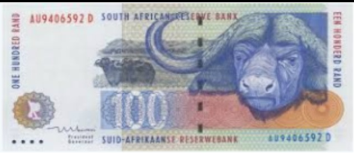<br>1-100 ZAR | 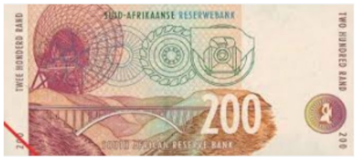<br>100-200 ZAR |
| Type of adherence support | 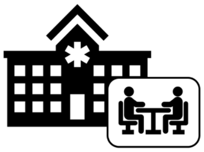<br>Counseling at clinic | 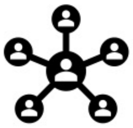<br>Adherence support groups |

### Choice Set 2



#### Which option would you choose?

| Characteristic | Option A | Option B |
| --- | --- | --- |
| Location of Service | Pharmacy image<br>Community setting | Community image<br>Pharmacy |
| The waiting time to get ART | Less than 1 hour | 2 hour maximum |
| The number of months of ART dispensed at a given time | x3<br>3 month | x1<br>1 month |
| Whether the provider is friendly or not | Provider image<br>Not friendly provider | Provider image<br>Friendly provider |
| The time of day you can get services | AM<br>Weekday morning (7:30am to 12pm) | Saturday Sunday<br>Weekends |
| How often visits are required (visit frequency) | x3<br>3 months | x6<br>6 months |
| The cost of getting to the service | FREE!<br>No cost | 1-100 ZAR |
| Type of adherence support | SMS/phone reminders | Adherence support groups |

### Choice Set 3

Block 2

#### Which option would you choose?

| Characteristic | Option A | Option B |
|---|---|---|
| Location of Service | Pharmacy image<br>Pharmacy | Community image<br>Community setting |
| The waiting time to get ART | 1 HOUR<br>Less than 1 hour | 1 HOUR 1 HOUR 1 HOUR<br>3 hour maximum |
| The number of months of ART dispensed at a given time | x6<br>6 month | x3<br>3 month |
| Whether the provider is friendly or not | Provider image<br>Friendly provider | Provider image<br>Not friendly provider |
| The time of day you can get services | Saturday Sunday<br>Weekends | AM<br>Weekday morning (7:30am to 12pm) |
| How often visits are required (visit frequency) | x1<br>1 month | x6<br>6 months |
| The cost of getting to the service | FREE!<br>No cost | 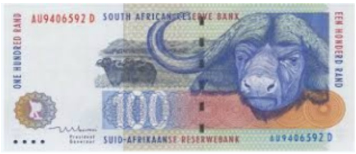<br>1-100 ZAR |
| Type of adherence support | SMS/phone reminders | Adherence support groups |

### Choice Set 4

Block 2

#### Which option would you choose?

| Characteristic | Option A | Option B |
|---|---|---|
| Location of Service | Pharmacy image<br>Clinic or hospital | Community image<br>Community setting |
| The waiting time to get ART | 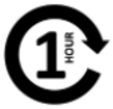<br>Less than 1 hour | 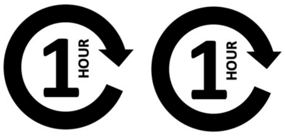<br>2 hour maximum |
| The number of months of ART dispensed at a given time | 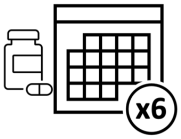<br>6 month | 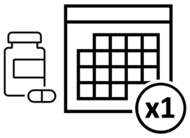<br>1 month |
| Whether the provider is friendly or not | Provider image<br>Not friendly provider | Provider image<br>Friendly provider |
| The time of day you can get services | 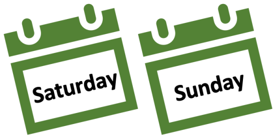<br>Weekends | 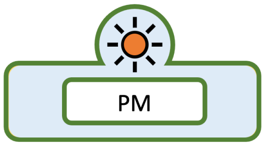<br>Weekday afternoon (2pm to 4pm) |
| How often visits are required (visit frequency) | 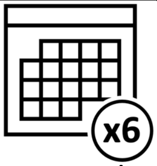<br>6 months | 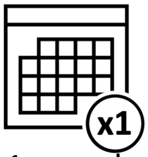<br>1 month |
| The cost of getting to the service | 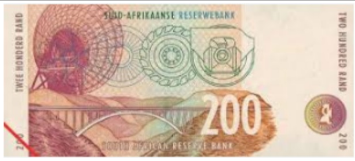<br>100-200 ZAR | 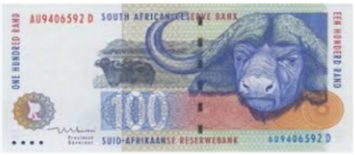<br>1-100 ZAR |
| Type of adherence support | 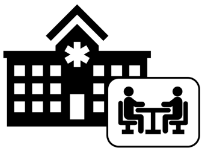<br>Counseling at clinic | 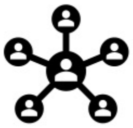<br>Adherence support groups |

### Choice Set 5

Block 2

Which option would you choose?

| Characteristic | Option A | Option B |
|---|---|---|
| Location of Service | Pharmacy image<br>Clinic or hospital | Community image<br>Pharmacy |
| The waiting time to get ART | 3 hour maximum | Less than 1 hour |
| The number of months of ART dispensed at a given time | x1<br>1 month | x6<br>6 month |
| Whether the provider is friendly or not | Provider image<br>Friendly provider | Provider image<br>Not friendly provider |
| The time of day you can get services | Saturday Sunday<br>Weekends | PM<br>Weekday afternoon (2pm to 4pm) |
| How often visits are required (visit frequency) | x6<br>6 months | x3<br>3 months |
| The cost of getting to the service | 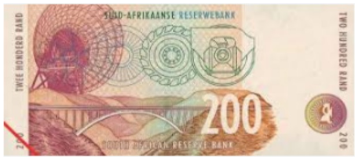<br>100-200 ZAR | FREE!<br>No cost |
| Type of adherence support | SMS/phone reminders | Adherence support groups |

### Choice Set 6



#### Which option would you choose?

| Characteristic | Option A | Option B |
|---|---|---|
| Location of Service | Pharmacy image<br>Pharmacy | Community image<br>Clinic or hospital |
| The waiting time to get ART | 3 hour maximum | Less than 1 hour |
| The number of months of ART dispensed at a given time | x3<br>3 month | x1<br>1 month |
| Whether the provider is friendly or not | Provider image<br>Not friendly provider | Provider image<br>Friendly provider |
| The time of day you can get services | Saturday Sunday<br>Weekends | AM<br>Weekday morning (7:30am to 12pm) |
| How often visits are required (visit frequency) | x1<br>1 month | x6<br>6 months |
| The cost of getting to the service | 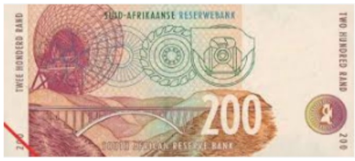<br>100-200 ZAR | FREE!<br>No cost |
| Type of adherence support | SMS/phone reminders | Counseling at clinic |

### Choice Set 7



Which option would you choose?

| Characteristic | Option A | Option B |
| --- | --- | --- |
| Location of Service | Pharmacy image<br>Clinic or hospital | Community image<br>Pharmacy |
| The waiting time to get ART | 2 hour maximum | Less than 1 hour |
| The number of months of ART dispensed at a given time | x6<br>6 month | x1<br>1 month |
| Whether the provider is friendly or not | Provider image<br>Friendly provider | Provider image<br>Not friendly provider |
| The time of day you can get services | Saturday Sunday<br>Weekends | AM<br>Weekday morning (7:30am to 12pm) |
| How often visits are required (visit frequency) | x3<br>3 months | x1<br>1 month |
| The cost of getting to the service | FREE!<br>No cost | 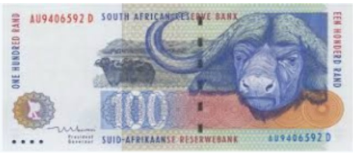<br>1-100 ZAR |
| Type of adherence support | Adherence support groups | Counseling at clinic |

### Choice Set 8

Block 2

Which option would you choose?

| Characteristic | Option A | Option B |
|---|---|---|
| Location of Service | Pharmacy image<br>Community setting | Community image<br>Pharmacy |
| The waiting time to get ART | 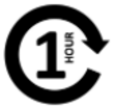<br>Less than 1 hour | 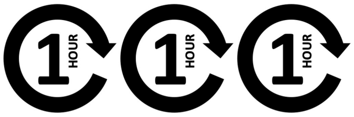<br>3 hour maximum |
| The number of months of ART dispensed at a given time | 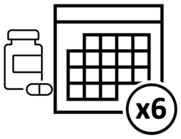<br>6 month | 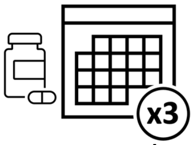<br>3 month |
| Whether the provider is friendly or not | Provider image<br>Not friendly provider | Provider image<br>Friendly provider |
| The time of day you can get services | 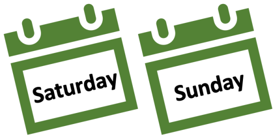<br>Weekends | 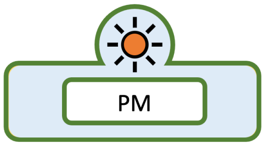<br>Weekday afternoon (2pm to 4pm) |
| How often visits are required (visit frequency) | 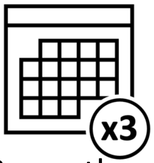<br>3 months | 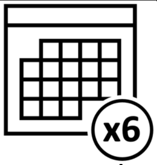<br>6 months |
| The cost of getting to the service | 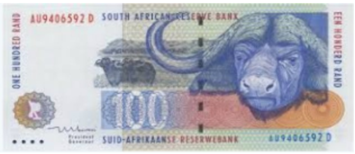<br>1-100 ZAR | FREE!<br>No cost |
| Type of adherence support | 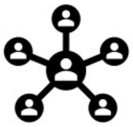<br>Adherence support groups | 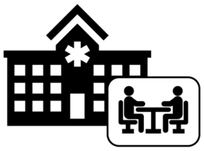<br>Counseling at clinic |

### Choice Set 9



#### Which option would you choose?

| Characteristic | Option A | Option B |
|---|---|---|
| Location of Service | Pharmacy image<br>Community setting | Community image<br>Clinic or hospital |
| The waiting time to get ART | 2 hour maximum | 3 hour maximum |
| The number of months of ART dispensed at a given time | x3<br>3 month | x1<br>1 month |
| Whether the provider is friendly or not | Provider image<br>Friendly provider | Provider image<br>Not friendly provider |
| The time of day you can get services | AM<br>Weekday morning (7:30am to 12pm) | PM<br>Weekday afternoon (2pm to 4pm) |
| How often visits are required (visit frequency) | x1<br>1 month | x6<br>6 months |
| The cost of getting to the service | 100-200 ZAR | FREE!<br>No cost |
| Type of adherence support | Counseling at clinic | SMS/phone reminders |

### Choice Set 1

Block 1

#### Which option would you choose?

| Characteristic | Option A | Option B |
|---|---|---|
| Location of Service | Pharmacy image<br>Clinic or hospital | Community image<br>Community setting |
| The waiting time to get ART | 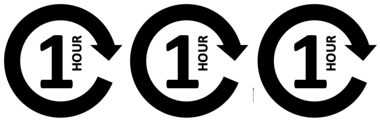<br>3 hour maximum | 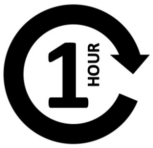<br>Less than 1 hour |
| The number of months of ART dispensed at a given time | 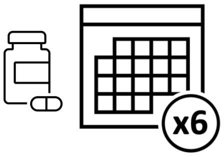<br>6 months of ART | 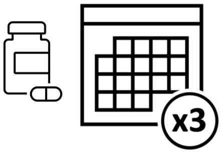<br>3 months of ART |
| Whether the provider is friendly or not | Provider image<br>Not friendly provider | Provider image<br>Friendly provider |
| The time of day you can get services | 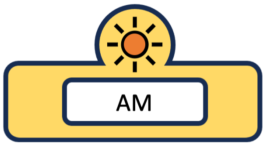<br>Weekday morning (7:30am to 12pm) | 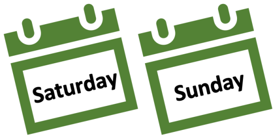<br>Weekends |
| How often visits are required (visit frequency for clinical visits) | 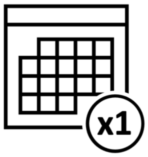<br>1 month | 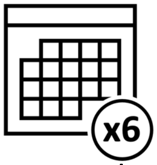<br>6 months |
| The cost of getting to the service (primarily transport costs) | 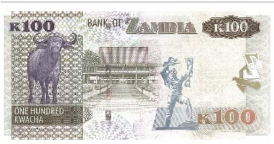<br>100+ ZMW | 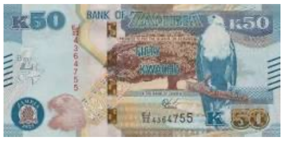<br>1-100 ZMW |
| Type of adherence support | 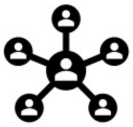<br>Adherence support groups | 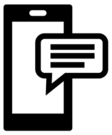<br>SMS/phone reminders |

### Choice Set 2

Block 1

#### Which option would you choose?

| Characteristic | Option A | Option B |
|---|---|---|
| Location of Service | Pharmacy image<br>Pharmacy | Community image<br>Clinic or hospital |
| The waiting time to get ART | 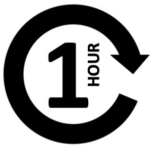<br>Less than 1 hour | 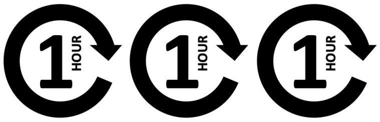<br>3 hour maximum |
| The number of months of ART dispensed at a given time | 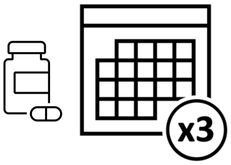<br>3 months of ART | 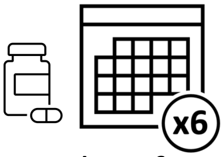<br>6 months of ART |
| Whether the provider is friendly or not | Provider image<br>Not friendly provider | Provider image<br>Friendly provider |
| The time of day you can get services | 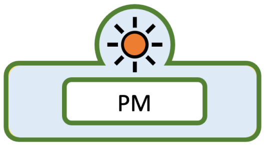<br>Weekday afternoon (2pm to 4pm) | 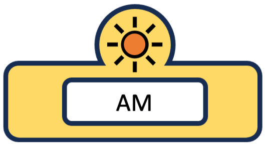<br>Weekday morning (7:30am to 12pm) |
| How often visits are required (visit frequency for clinical visits) | 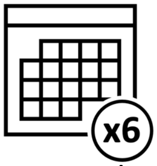<br>6 months | 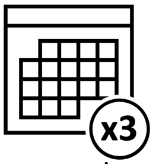<br>3 months |
| The cost of getting to the service (primarily transport costs) | 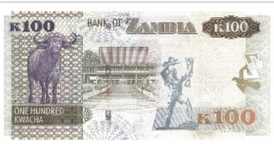<br>100+ ZMW | 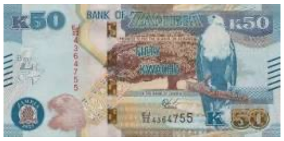<br>1-100 ZMW |
| Type of adherence support | 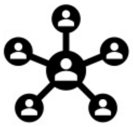<br>Adherence support groups | 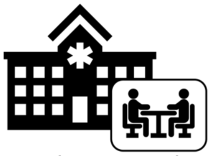<br>Counseling at clinic |

### Choice Set 3

Block 1

Which option would you choose?

| Characteristic | Option A | Option B |
|---|---|---|
| Location of Service | Pharmacy image<br>Community setting | Community image<br>Pharmacy |
| The waiting time to get ART | 3 hour maximum | 2 hour maximum |
| The number of months of ART dispensed at a given time | x3<br>3 months of ART | x6<br>6 months of ART |
| Whether the provider is friendly or not | Provider image<br>Friendly provider | Provider image<br>Not friendly provider |
| The time of day you can get services | Saturday Sunday<br>Weekends | AM<br>Weekday morning (7:30am to 12pm) |
| How often visits are required (visit frequency for clinical visits) | x1<br>1 month | x6<br>6 months |
| The cost of getting to the service (primarily transport costs) | FREE!<br>No cost | 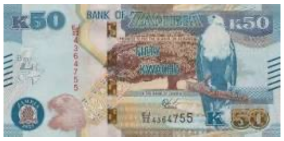<br>1-100 ZMW |
| Type of adherence support | Counseling at clinic | SMS/phone reminders |

*Zambia DCE*
*Block 1*

### Choice Set 4

**Block 1**

#### Which option would you choose?

| Characteristic | Option A | Option B |
|---|---|---|
| Location of Service | Pharmacy image<br>Clinic or hospital | Community image<br>Community setting |
| The waiting time to get ART | 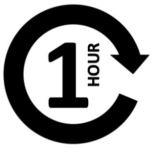<br>Less than 1 hour | 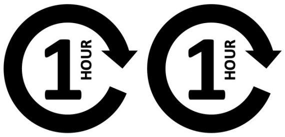<br>2 hour maximum |
| The number of months of ART dispensed at a given time | 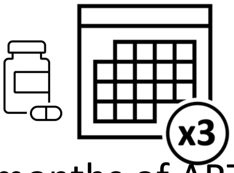<br>3 months of ART | 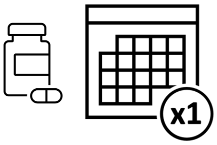<br>1 month of ART |
| Whether the provider is friendly or not | Provider image<br>Friendly provider | Provider image<br>Not friendly provider |
| The time of day you can get services | 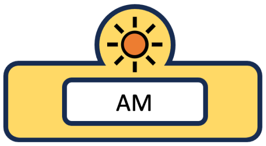<br>Weekday morning (7:30am to 12pm) | 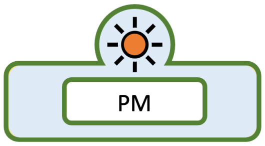<br>Weekday afternoon (2pm to 4pm) |
| How often visits are required (visit frequency for clinical visits) | 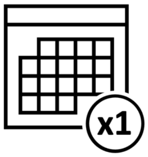<br>1 month | 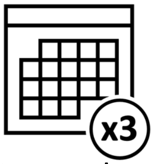<br>3 months |
| The cost of getting to the service (primarily transport costs) | 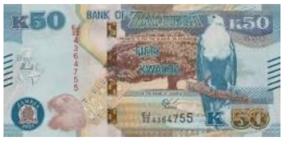<br>1-100 ZMW | 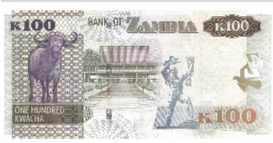<br>100+ ZMW |
| Type of adherence support | 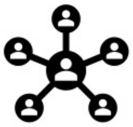<br>Adherence support groups | 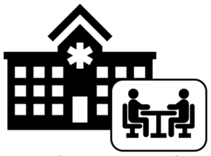<br>Counseling at clinic |

### Choice Set 5



Which option would you choose?

| Characteristic | Option A | Option B |
|---|---|---|
| Location of Service | Pharmacy image<br>Pharmacy | Community image<br>Clinic or hospital |
| The waiting time to get ART | 3 hour maximum | 2 hour maximum |
| The number of months of ART dispensed at a given time | 6 months of ART | 1 month of ART |
| Whether the provider is friendly or not | Provider image<br>Friendly provider | Provider image<br>Not friendly provider |
| The time of day you can get services | Weekday afternoon (2pm to 4pm) | Weekday morning (7:30am to 12pm) |
| How often visits are required (visit frequency for clinical visits) | 3 months | 1 month |
| The cost of getting to the service (primarily transport costs) | 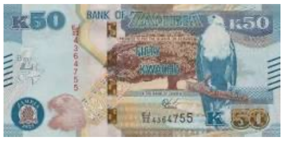<br>1-100 ZMW | FREE!<br>No cost |
| Type of adherence support | Counseling at clinic | Adherence support groups |

### Choice Set 6

Block 1

Which option would you choose?

| Characteristic | Option A | Option B |
|---|---|---|
| Location of Service | Pharmacy image<br>Pharmacy | Community image<br>Clinic or hospital |
| The waiting time to get ART | 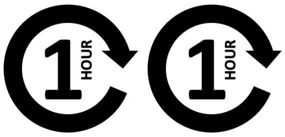<br>2 hour maximum | 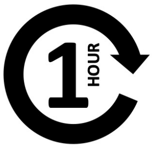<br>Less than 1 hour |
| The number of months of ART dispensed at a given time | 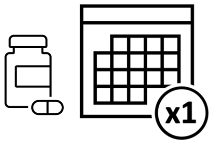<br>1 month of ART | 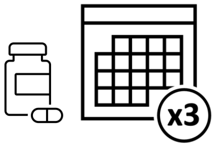<br>3 months of ART |
| Whether the provider is friendly or not | Provider image<br>Not friendly provider | Provider image<br>Friendly provider |
| The time of day you can get services | 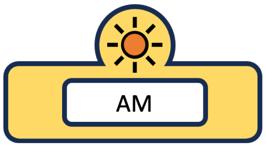<br>Weekday morning (7:30am to 12pm) | 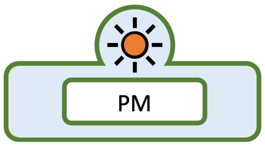<br>Weekday afternoon (2pm to 4pm) |
| How often visits are required (visit frequency for clinical visits) | 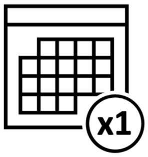<br>1 month | 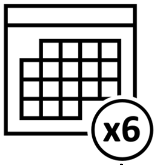<br>6 months |
| The cost of getting to the service (primarily transport costs) | FREE!<br>No cost | 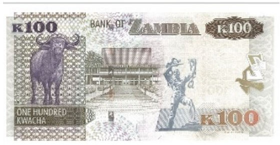<br>100+ ZMW |
| Type of adherence support | 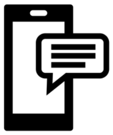<br>SMS/phone reminders | 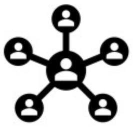<br>Adherence support groups |

### Choice Set 7

Block 1

#### Which option would you choose?

| Characteristic | Option A | Option B |
|---|---|---|
| Location of Service | Pharmacy image<br>Clinic or hospital | Community image<br>Community setting |
| The waiting time to get ART | 1 HOUR 1 HOUR<br>2 hour maximum | 1 HOUR 1 HOUR 1 HOUR<br>3 hour maximum |
| The number of months of ART dispensed at a given time | x3<br>3 months of ART | x6<br>6 months of ART |
| Whether the provider is friendly or not | Provider image<br>Friendly provider | Provider image<br>Not friendly provider |
| The time of day you can get services | PM<br>Weekday afternoon (2pm to 4pm) | Saturday Sunday<br>Weekends |
| How often visits are required (visit frequency for clinical visits) | x3<br>3 months | x6<br>6 months |
| The cost of getting to the service (primarily transport costs) | 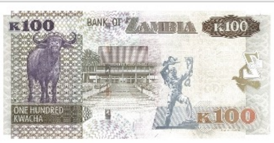 <br>100+ ZMW | FREE!<br>No cost |
| Type of adherence support | SMS/phone reminders | Adherence support groups |

Zambia DCE
Block 1

### Choice Set 8

Block 1

Which option would you choose?

| Characteristic | Option A | Option B |
|---|---|---|
| Location of Service | Pharmacy image<br>Community setting | Community image<br>Clinic or hospital |
| The waiting time to get ART | 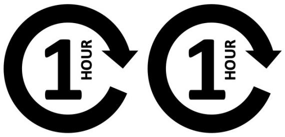<br>2 hour maximum | 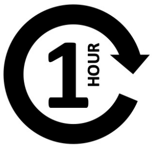<br>Less than 1 hour |
| The number of months of ART dispensed at a given time | 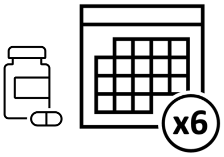<br>6 months of ART | 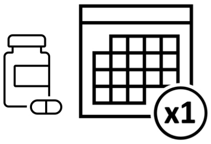<br>1 month of ART |
| Whether the provider is friendly or not | Provider image<br>Friendly provider | Provider image<br>Not friendly provider |
| The time of day you can get services | 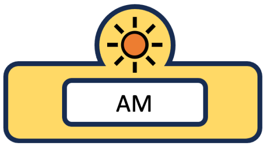<br>Weekday morning (7:30am to 12pm) | 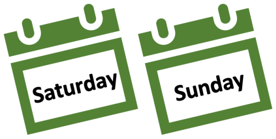<br>Weekends |
| How often visits are required (visit frequency for clinical visits) | 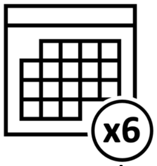<br>6 months | 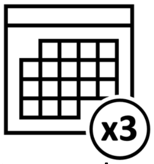<br>3 months |
| The cost of getting to the service (primarily transport costs) | 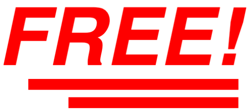<br>No cost | 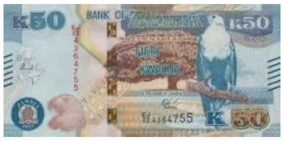<br>1-100 ZMW |
| Type of adherence support | 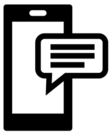<br>SMS/phone reminders | 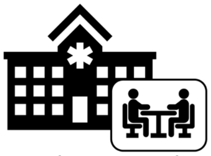<br>Counseling at clinic |

### Choice Set 9

Block 1

#### Which option would you choose?

| Characteristic | Option A | Option B |
|---|---|---|
| Location of Service | Pharmacy image<br>Pharmacy | Community image<br>Community setting |
| The waiting time to get ART | 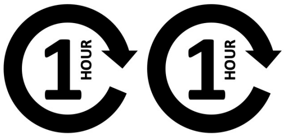<br>2 hour maximum | 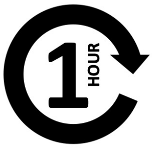<br>Less than 1 hour |
| The number of months of ART dispensed at a given time | 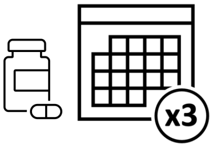<br>3 months of ART | 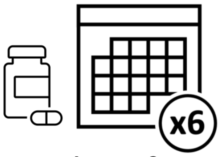<br>6 months of ART |
| Whether the provider is friendly or not | Provider image<br>Not friendly provider | Provider image<br>Friendly provider |
| The time of day you can get services | 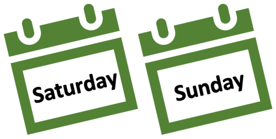<br>Weekends | 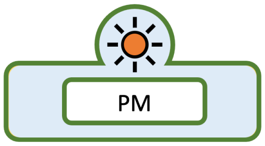<br>Weekday afternoon (2pm to 4pm) |
| How often visits are required (visit frequency for clinical visits) | 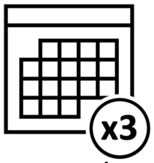<br>3 months | 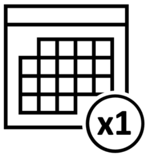<br>1 month |
| The cost of getting to the service (primarily transport costs) | 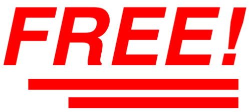<br>No cost | 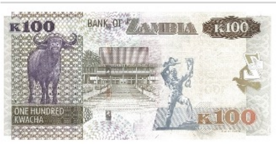<br>100+ ZMW |
| Type of adherence support | 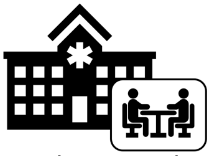<br>Counseling at clinic | 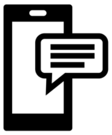<br>SMS/phone reminders |

### Choice Set 1

Block 2

#### Which option would you choose?

| Characteristic | Option A | Option B |
|---|---|---|
| Location of Service | Pharmacy image<br>Clinic or hospital | Community image<br>Pharmacy |
| The waiting time to get ART | 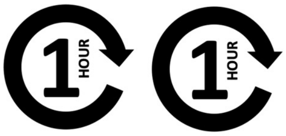 <br>2 hour maximum | 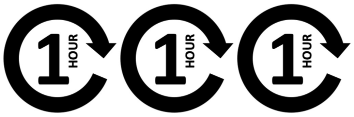 <br>3 hour maximum |
| The number of months of ART dispensed at a given time | 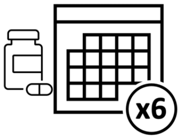 <br>6 month | 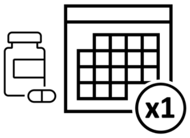 <br>1 month |
| Whether the provider is friendly or not | Provider image<br>Not friendly provider | Provider image<br>Friendly provider |
| The time of day you can get services | 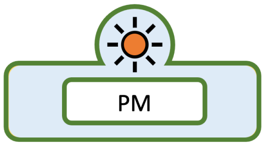 <br>Weekday afternoon (2pm to 4pm) | 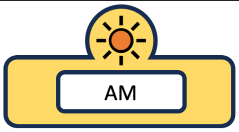 <br>Weekday morning (7:30am to 12pm) |
| How often visits are required (visit frequency for clinical visits) | 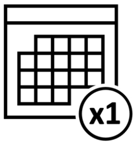 <br>1 month | 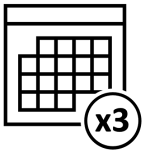 <br>3 months |
| The cost of getting to the service (primarily transport costs) | 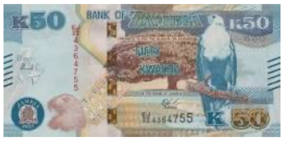<br>1-100 ZMW | 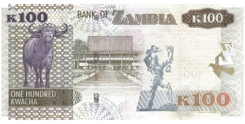<br>100+ ZMW |
| Type of adherence support | 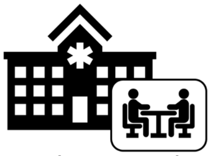<br>Counseling at clinic | 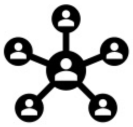<br>Adherence support groups |

### Choice Set 2

Block 2

#### Which option would you choose?

| Characteristic | Option A | Option B |
|---|---|---|
| Location of Service | Pharmacy image<br>Community setting | Community image<br>Pharmacy |
| The waiting time to get ART | 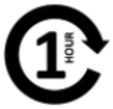<br>Less than 1 hour | 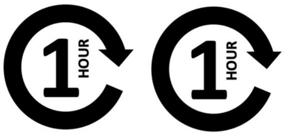<br>2 hour maximum |
| The number of months of ART dispensed at a given time | 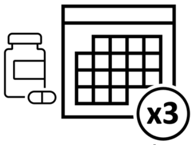<br>3 month | 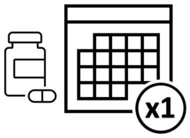<br>1 month |
| Whether the provider is friendly or not | Provider image<br>Not friendly provider | Provider image<br>Friendly provider |
| The time of day you can get services | 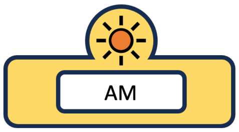<br>Weekday morning (7:30am to 12pm) | 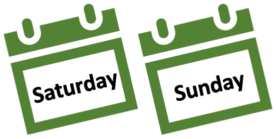<br>Weekends |
| How often visits are required (visit frequency) | 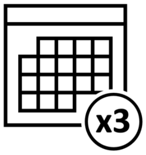<br>3 months | 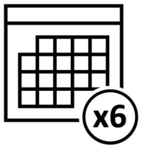<br>6 months |
| The cost of getting to the service | 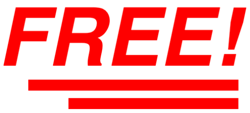<br>No cost | 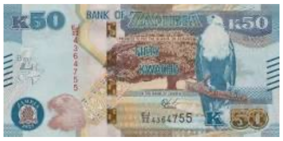<br>1-100 ZMW |
| Type of adherence support | 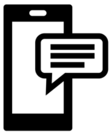<br>SMS/phone reminders | 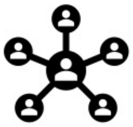<br>Adherence support groups |

### Choice Set 3



#### Which option would you choose?

| Characteristic | Option A | Option B |
|---|---|---|
| Location of Service | Pharmacy image<br>Pharmacy | Community image<br>Community setting |
| The waiting time to get ART | 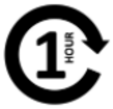<br>Less than 1 hour | 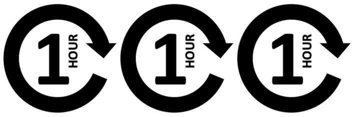<br>3 hour maximum |
| The number of months of ART dispensed at a given time | 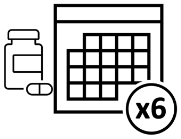<br>6 month | 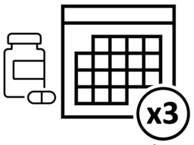<br>3 month |
| Whether the provider is friendly or not | Provider image<br>Friendly provider | Provider image<br>Not friendly provider |
| The time of day you can get services | 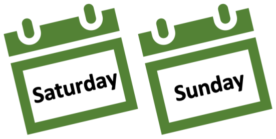<br>Weekends | 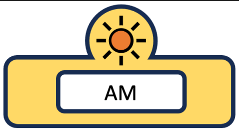<br>Weekday morning (7:30am to 12pm) |
| How often visits are required (visit frequency for clinical visits) | 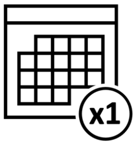<br>1 month | 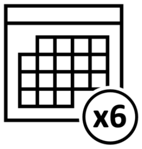<br>6 months |
| The cost of getting to the service (primarily transport costs) | FREE!<br>No cost | 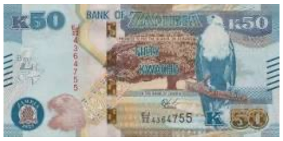<br>1-100 ZMW |
| Type of adherence support | 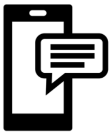<br>SMS/phone reminders | 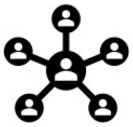<br>Adherence support groups |

*Zambia DCE*
*Block 2*

### Choice Set 4

Block 2

#### Which option would you choose?

| Characteristic | Option A | Option B |
|---|---|---|
| Location of Service | Pharmacy image<br>Clinic or hospital | Community image<br>Community setting |
| The waiting time to get ART | 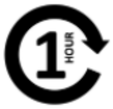 <br>Less than 1 hour | 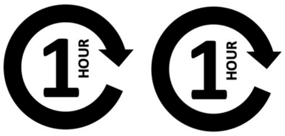 <br>2 hour maximum |
| The number of months of ART dispensed at a given time | 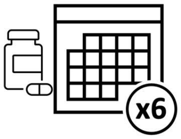 <br>6 month | 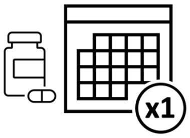 <br>1 month |
| Whether the provider is friendly or not | Provider image<br>Not friendly provider | Provider image<br>Friendly provider |
| The time of day you can get services | 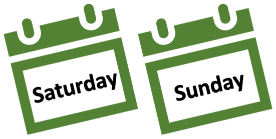 <br>Weekends | 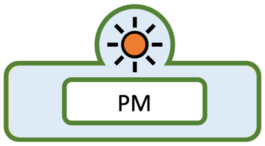 <br>Weekday afternoon (2pm to 4pm) |
| How often visits are required (visit frequency) | 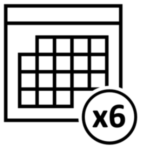 <br>6 months | 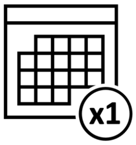 <br>1 month |
| The cost of getting to the service | 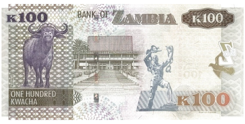 <br>100+ ZMW | 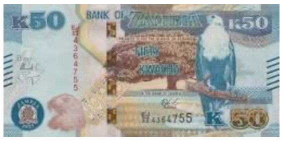 <br>1-100 ZMW |
| Type of adherence support | 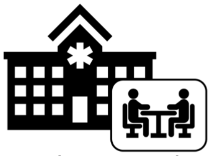<br>Counseling at clinic | 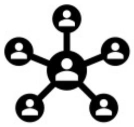<br>Adherence support groups |

### Choice Set 5

Block 2

#### Which option would you choose?

| Characteristic | Option A | Option B |
|---|---|---|
| Location of Service | Pharmacy image<br>Clinic or hospital | Community image<br>Pharmacy |
| The waiting time to get ART | 3 hour maximum | Less than 1 hour |
| The number of months of ART dispensed at a given time | x1<br>1 month | x6<br>6 month |
| Whether the provider is friendly or not | Provider image<br>Friendly provider | Provider image<br>Not friendly provider |
| The time of day you can get services | Saturday Sunday<br>Weekends | PM<br>Weekday afternoon (2pm to 4pm) |
| How often visits are required (visit frequency for clinical visits) | x6<br>6 months | x3<br>3 months |
| The cost of getting to the service (primarily transport costs) | 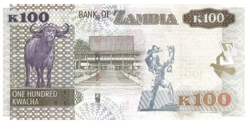<br>100+ ZMW | FREE!<br>No cost |
| Type of adherence support | SMS/phone reminders | Adherence support groups |

### Choice Set 6

Block 2

Which option would you choose?

| Characteristic | Option A | Option B |
|---|---|---|
| Location of Service | Pharmacy image<br>Pharmacy | Community image<br>Clinic or hospital |
| The waiting time to get ART | 3 hour maximum | Less than 1 hour |
| The number of months of ART dispensed at a given time | x3<br>3 month | x1<br>1 month |
| Whether the provider is friendly or not | Provider image<br>Not friendly provider | Provider image<br>Friendly provider |
| The time of day you can get services | Saturday Sunday<br>Weekends | AM<br>Weekday morning (7:30am to 12pm) |
| How often visits are required (visit frequency for clinical visits) | x1<br>1 month | x6<br>6 months |
| The cost of getting to the service (primarily transport costs) | 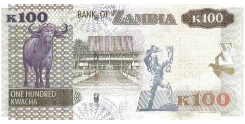<br>100+ ZMW | FREE!<br>No cost |
| Type of adherence support | SMS/phone reminders | Counseling at clinic |

### Choice Set 7

Block 2

#### Which option would you choose?

| Characteristic | Option A | Option B |
|---|---|---|
| Location of Service | Pharmacy image<br>Clinic or hospital | Community image<br>Pharmacy |
| The waiting time to get ART | 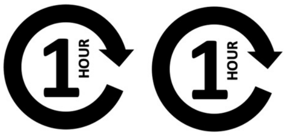 <br>2 hour maximum | 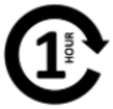 <br>Less than 1 hour |
| The number of months of ART dispensed at a given time | 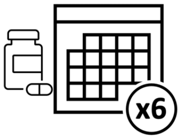 <br>6 month | 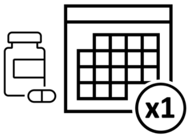 <br>1 month |
| Whether the provider is friendly or not | Provider image<br>Friendly provider | Provider image<br>Not friendly provider |
| The time of day you can get services | 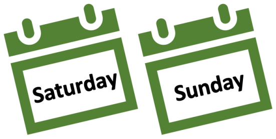 <br>Weekends | 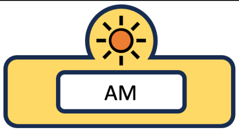 <br>Weekday morning (7:30am to 12pm) |
| How often visits are required (visit frequency for clinical visits) | 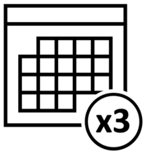 <br>3 months | 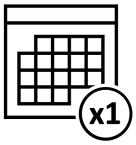 <br>1 month |
| The cost of getting to the service (primarily transport costs) | 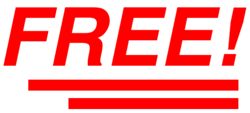 <br>No cost | 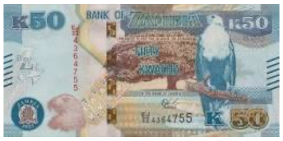<br>1-100 ZMW |
| Type of adherence support | 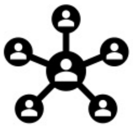<br>Adherence support groups | 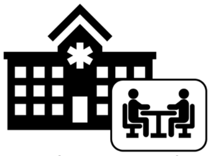<br>Counseling at clinic |

### Choice Set 8
Which option would you choose?



| Characteristic | Option A | Option B |
|---|---|---|
| Location of Service | Pharmacy image<br>Community setting | Community image<br>Pharmacy |
| The waiting time to get ART | 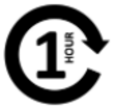<br>Less than 1 hour | 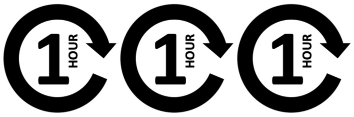<br>3 hour maximum |
| The number of months of ART dispensed at a given time | 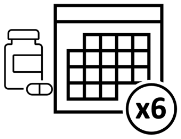<br>6 month | 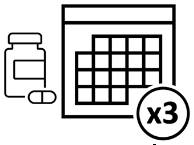<br>3 month |
| Whether the provider is friendly or not | Provider image<br>Not friendly provider | Provider image<br>Friendly provider |
| The time of day you can get services | 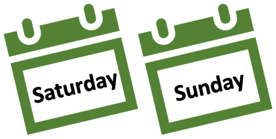<br>Weekends | 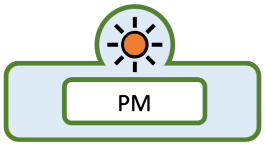<br>Weekday afternoon (2pm to 4pm) |
| How often visits are required (visit frequency for clinical visits) | 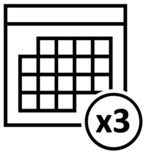<br>3 months | 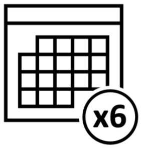<br>6 months |
| The cost of getting to the service (primarily transport costs) | 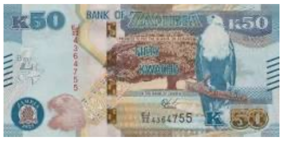<br>1-100 ZMW | FREE!<br>No cost |
| Type of adherence support | 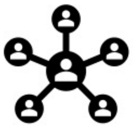<br>Adherence support groups | 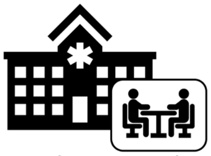<br>Counseling at clinic |

### Choice Set 9



Which option would you choose?

| Characteristic | Option A | Option B |
|---|---|---|
| Location of Service | Pharmacy image<br>Community setting | Community image<br>Clinic or hospital |
| The waiting time to get ART | 1 HOUR 1 HOUR<br>2 hour maximum | 1 HOUR 1 HOUR 1 HOUR<br>3 hour maximum |
| The number of months of ART dispensed at a given time | x3<br>3 month | x1<br>1 month |
| Whether the provider is friendly or not | Provider image<br>Friendly provider | Provider image<br>Not friendly provider |
| The time of day you can get services | AM<br>Weekday morning (7:30am to 12pm) | PM<br>Weekday afternoon (2pm to 4pm) |
| How often visits are required (visit frequency for clinical visits) | x1<br>1 month | x6<br>6 months |
| The cost of getting to the service (primarily transport costs) | 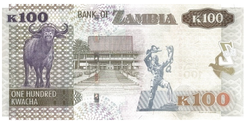<br>100+ ZMW | FREE!<br>No cost |
| Type of adherence support | Counseling at clinic | SMS/phone reminders |
