## Supplementary File 2. Variable definitions for "Client preferences for service delivery during the early treatment period in South Africa and Zambia: Mixed-method findings from a discrete choice experiment and concurrent focus group discussions"

Supplementary Table 2. Variable definitions used for conditional logit model

| Variable Name | Definition |
| --- | --- |
| prefer_studyID | Unique PREFER Study ID |
| obsid | Represents each unique choice made in the dataset (required by Stata) |
| alt | Alternative within each choice set. Each choice task had two choice sets, and alt distinguishes between them |
| cno | Represents the choice number in each choice task. Each respondent made 9 choices, so this variable ranges between 1-9 and repeats for each participant |
| choiceset | Represents the choice number in the DCE. The activity is made up of 18 total choices, so this variable ranges between 1-18. |
| location_clinic | Dummy variable for  Attribute: Location of service  Level: Clinic or hospital |
| location_pharm | Dummy variable for  Attribute: Location of service  Level: Pharmacy |
| location_comm | Dummy variable for  Attribute: Location of service  Level: Community |
| wait_0 | Dummy variable for  Attribute: Waiting time at facilities  Level: Less than 1 hour |
| wait_1 | Dummy variable for  Attribute: Waiting time at facilities  Level: 2 hour maximum |
| wait_2 | Dummy variable for  Attribute: Waiting time at facilities  Level: 3 hour maximum |
| dispense_0 | Dummy variable for  Attribute: Dispensing interval  Level: 1 month |
| dispense_1 | Dummy variable for  Attribute: Dispensing interval  Level: 2 months |
| dispense_2 | Dummy variable for  Attribute: Dispensing interval  Level: 3 months |
| friendly | Variable for  Attribute: Provider disposition  Level 0: Unfriendly  Level 1: Friendly |
| time_morning | Dummy variable for  Attribute: The time of day you can get services  Level: Weekday morning |
| time_afternoon | Dummy variable for  Attribute: The time of day you can get services  Level: Weekday afternoon |
| time_weekend | Dummy variable for,  Attribute: The time of day you can get services  Level: Weekend |
| freq_0 | Dummy variable for,  Attribute: Visit frequency  Level: 1 month |
| freq_1 | Dummy variable for,  Attribute: Visit frequency  Level: 3 months |
| freq_2 | Dummy variable for,  Attribute: Visit frequency  Level: 6 months |
| cost_0 | Dummy variable for,  Attribute: Cost of obtaining service  Level: Free |
| cost_1 | Dummy variable for,  Attribute: Cost of obtaining service  Level: 1-100 ZAR/ZMW |
| cost_2 | Dummy variable for,  Attribute: Cost of obtaining service  Level: >100 ZAR/ZMW |
| type_counsel | Dummy variable for,  Attribute: Type of adherence support  Level: Counseling at clinic |
| type_phone | Dummy variable for,  Attribute: Type of adherence support  Level: SMS or phone reminders |
| type_asg | Dummy variable for,  Attribute: Type of adherence support  Level: Adherence support groups |
| choice | Represents respondent’s choice for a given choice set |
