## Supplementary Table 1. Full PREFER demographic characteristics for "Client preferences for service delivery during the early treatment period in South Africa and Zambia: Mixed-method findings from a discrete choice experiment and concurrent focus group discussions"

| **Characteristic, n (%)** | **South Africa** | | **Zambia** | |
| --- | --- | --- | --- | --- |
|  | **DCE/FGD** | **PREFER Baseline Survey** | **DCE/FGD** | **PREFER Baseline Survey** |
|  | **(N=123)** | **(N=1098)** | **(N=126)** | **(N=771)** |
| Female | 103 (84) | 786 (72) | 70 (56) | 514 (67) |
| Age, median (IQR) | 33 (28, 41) | 33 (27, 41) | 35 (30, 44) | 32 (27, 40) |
| Education |  |  |  |  |
| *Primary or less* | 39 (32) | 409 (37) | 59 (47) | 386 (50) |
| *Secondary* | 66 (54) | 525 (48) | 58 (46) | 329 (43) |
| *Post-secondary* | 18 (14) | 164 (15) | 9 (7) | 56 (7) |
| Employment status |  |  |  |  |
| *Formal employment* | 15 (12) | 240 (22) | 12 (10) | 73 (9) |
| *Informal employment* | 24 (19) | 216 (20) | 65 (52) | 413 (54) |
| *Unemployed* | 74 (60) | 562 (51) | 47 (37) | 267 (35) |
| *Student/trainee* | 10 (8) | 80 (7) | 2 (2) | 18 (2) |
| Time on ART at PREFER survey enrollment |  |  |  |  |
| *Day of survey* | 43 (35) | 419 (38) | 40 (32) | 261 (34) |
| *0-180 days* | 80 (65) | 679 (62) | 86 (68) | 510 (66) |
| Would find it difficult/very difficult procuring 100 Rands if needed for medical treatment | 76 (61) | 615 (56) | 111 (88) | 629 (82) |

**Includes clients newly initiating or re-initiating ART on the day of the PREFER Baseline Survey*
